# A microsimulation-based framework for mitigating societal bias in primary care data

**DOI:** 10.1101/2025.04.03.25325206

**Authors:** Agata Foryciarz, Fernando Alarid-Escudero, Gabriela Basel, Marika Cusick, Robert L. Phillips, Andrew Bazemore, Alyce Adams, Sherri Rose

## Abstract

**Purpose:** The data generating mechanisms underlying health care data are infrequently considered, leading to inequitable equilibria being reinforced throughout the care continuum. As race-based criteria are reassessed, the effect of those criteria on patterns of disease progression should also be reevaluated. We proposed a novel microsimulation-based framework for attenuating societal bias in primary care registry data to study this.

**Methods:** Our data transformation framework enables generating counterfactual outcome distributions that would have been observed in the absence of race-based diagnosis and treatment criteria. We developed a continuous-time, discrete-event individual-level simulation model of kidney function decline, measured by estimated glomerular filtration rate (eGFR). The model simulates individual eGFR trajectories over time. eGFR decline is accelerated by hypertension, diabetes, and reaching chronic kidney disease stage 3a, and can be delayed by interventions, which are applied based on eGFR level, measured with or without an adjustment for Black race. A Bayesian calibration procedure was applied to identify rates of eGFR decline corresponding to stage distributions in the cohort.

**Results:** Under the counterfactual scenario without a race adjustment, Black individuals qualify for diagnosis earlier, and non-Black individuals later, than under the reference scenario with race adjustment. The difference was largest for earlier stages and smaller at each consecutive stage. We do not observe differences in life expectancy between the two scenarios.

**Limitations:** Large variability in the prevalence of treatment and heterogeneity in treatment effectiveness may impact our results.

**Conclusions:** Our data transformation framework demonstrates how the explicit representation of the data generation process could inform the effect of policy changes on clinical data distributions. The framework can flexibly be adapted to mitigate bias in other health data.

**Highlights:**

- We developed a novel data transformation framework for attenuating societal biases in data using microsimulation models in a study of chronic kidney disease progression with primary care data.
- The removal of race-based diagnostic criteria in our simulations changed the timing of qualification for chronic kidney disease diagnosis, ranging from 0.6 years to 9.6 years, with opposite effects for Black and non-Black patients.
- The simulated differences in expected survival after removing the race adjustment did not exceed 2 months among individuals who developed CKD.
- The explicit representation of the data generation process can help anticipate the effect that policy changes can have on clinical data distributions.

## 1 Introduction

Primary care plays a central role in the management of chronic disease and has the potential to address factors early in disease progression that have downstream health effects. However, there are substantial disparities in the health system that contribute to persistently worse health outcomes for minoritized groups.^1,2^ New, large primary care datasets may better inform interventions that ameliorate health inequities as well as statistical tools that encourage earlier diagnosis and treatment for patients who have experienced delayed care. However, these data reflect the existing inequitable equilibrium of the healthcare system, as they are encoded with societal biases, including racism, race-based treatment criteria, access disparities, and unmeasured differential exposure to social factors.^3–5^ This can lead to lower-quality evidence for groups underrepresented in health records due to access barriers, and incorrect statistical inferences when confounders, such as social exposures and race-associated under-treatment patterns, are not appropriately accounted for. Hence, there is a risk that using these data as they are to build statistical tools will further perpetuate biases.^6^ Additionally, healthcare data rarely incorporate information on social drivers of health, social mechanisms and structures beyond the healthcare system that impact health, and are among the most important contributors to health inequities.^7^

Approaches for transforming data to mitigate societal bias, referred to elsewhere as data de-biasing, have previously been proposed in the algorithmic fairness literature.^8–10^ These approaches assume that the data encode a form of societal bias, which arises from a socially biased data generation process, measurement error, or unmeasured confounders. They include re-labeling, resampling or reweighing data, or generating intermediate data representations where some of the information and correlation structure is removed. These methods typically try to change data *as if* the process generating the data was different, but do not usually formally define or explicitly model the change. They also do not typically incorporate social drivers of health, which contribute to the data generation process.^11^

In this work, we develop a new framework for data transformations that explicitly encodes changes to the biased data generating process to reflect a desired equilibrium. Our proposed method is a novel pre-processing algorithmic fairness approach that utilizes microsimulation modeling to mechanistically define the data generating process and generate new values under clearly defined changes to the data generating process. The simulated data can then be used to inform future interventions and policy decisions. To our knowledge, this is the first microsimulation-based data transformation approach.

To illustrate our framework, we study chronic kidney disease (CKD), a heterogeneous, progressive condition affecting 1 in 7 Americans.^12^ Early diagnosis and treatment are crucial for maintaining health and preventing irreversible damage. The condition is classified into six stages corresponding to an increasing degree of kidney damage. Stages 1 and 2 are often asymptomatic and diagnosis requires urine tests for albuminuria, while the remaining stages (3a, 3b, 4, and 5) can be identified using only the estimated glomerular filtration rate (eGFR), which corresponds to the percentage of remaining kidney function.^13^ Appropriate management of CKD differs depending on disease etiology, comorbidities, and progression speed.^14^ In Stage 5, also referred to as end-stage renal disease (ESRD), the only treatment options are dialysis or kidney transplant. Timely primary and specialist care has been associated with reductions in the yearly rate of eGFR decline and reduced mortality.^15–22^ In the U.S., CKD is more prevalent among racial and ethnic minorities than in white patients.^23^ Black patients have higher rates of ESRD and faster progression through CKD stages compared to white patients, despite similar rates of CKD diagnosis between the two groups.^24^ A range of social and structural factors contributes to those inequities,^25–27^ including adverse environmental exposures and neighborhood conditions, as well as suboptimal care patterns.^28,29^

Race-adjusted formulas for estimating eGFR have been used for diagnosis and treatment decisions for decades.^30–36^ This race adjustment for Black patients has faced significant criticism for lack of clear biological justification and perpetuating racial bias.^38^ The implementation of race adjustment in the eGFR formula likely contributed to delayed CKD diagnosis and treatment for Black patients as well as faster disease progression and higher mortality since it overestimated their eGFR, assigning them to less severe CKD categories.^24,34,38–41^ In 2021, a new eGFR equation without race adjustment was proposed, with uptake by the majority of U.S. labs by 2023.^39,42,43^ It has been hypothesized that using the 2021 formula may reduce delays in the treatment of Black patients by encouraging earlier initiation of stage-specific treatment and care.^39^

We use our data transformation framework to simulate CKD trajectories. These trajectories correspond to primary care data that may have been observed under a more equitable data generation process if the current eGFR criteria (without race adjustment) had been in effect since 2017. We will explicitly account for changes in the timing of diagnosis and stage assignment, as well as changes in CKD progression and mortality resulting from updating a 2009 formula to the 2021 formula, which leads to changes in stage-specific interventions. Additionally, we explicitly model social drivers of health in the data generating process.

## 2 Methods

### 2.1 General framework

We propose the following mechanism of how primary care datasets come to reflect the inequitable equilibrium of the healthcare system, as illustrated in Figure 1. In this general framework, individual characteristics (such as gender, socioeconomic status, race and ethnicity), together with geographic location, impact one’s exposure to social drivers of health as well as the access and quality of medical care received. All of these factors then impact individual health trajectories over time. This encompasses a number of known mechanisms generating health inequities, including differential exposure and vulnerability to psychosocial and material exposure by social status, sociodemographic differences in access to care and regional differences in quality of care, as well as race-based treatment criteria.^38,44^ This framework captures a normative concept of societal bias, as opposed to statistical bias (e.g., sampling bias and measurement error), which can additionally be introduced when health trajectories then get recorded as medical data^45,46^.

**Figure 1:**
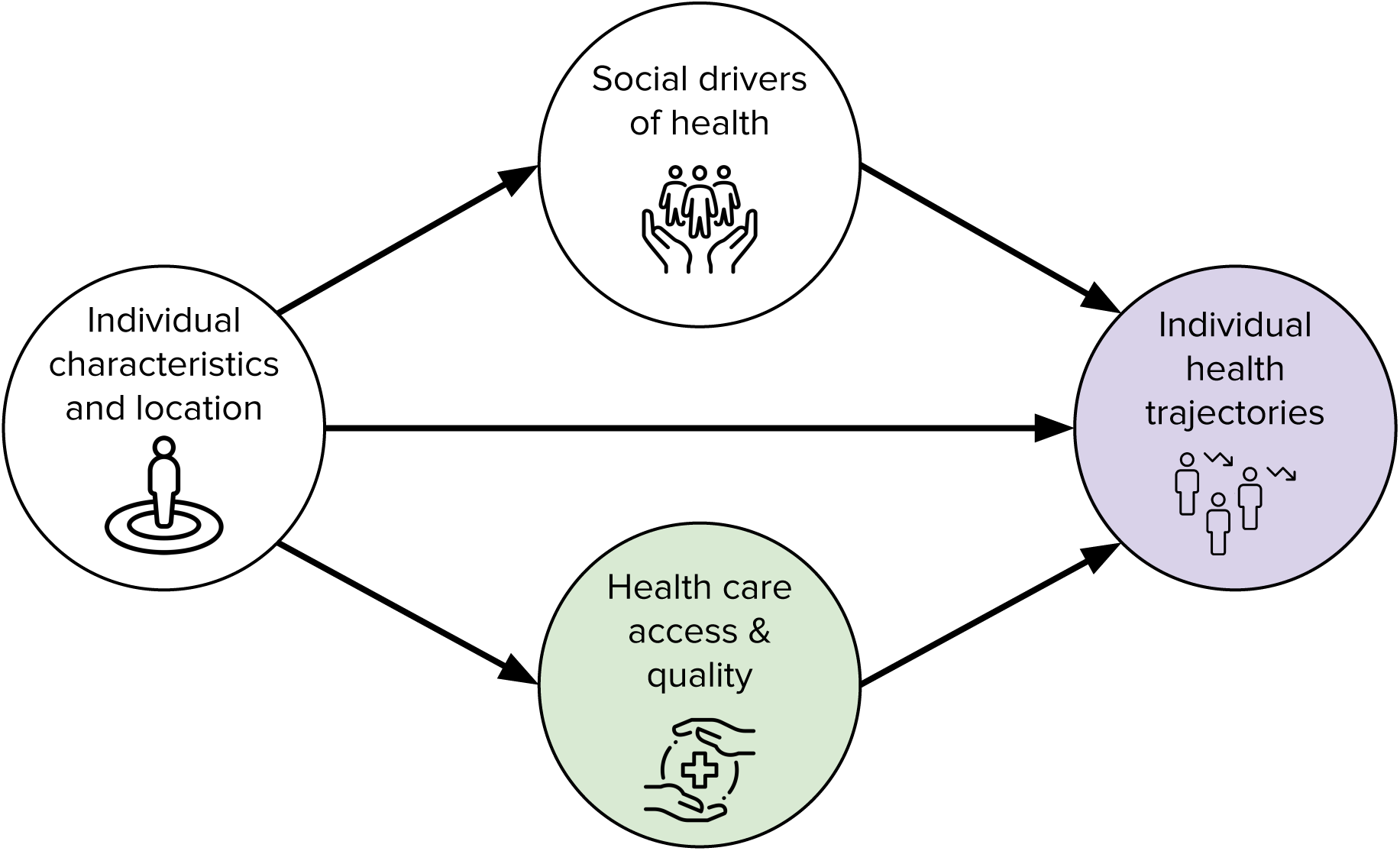
The proposed data bias mitigation framework, with intervention node (green) and outcome node (purple) marked. Societal bias arises from the association between individual characteristics and health care access and quality, as well as social drivers of health. By applying interventions that change health care access and quality, the model is able to simulate resulting changes in the distribution of simulated health trajectories.

Our framework allows us to generate realistic data simulations under various counterfactual scenarios, such as those corresponding to changes in social policies or medical guidelines. This could include changes in the timing of diagnosis or treatment as well as changes in exposure to risk factors that directly affect health. This approach constitutes an innovation relative to existing data bias mitigation strategies proposed in the literature by generating data modifications corresponding to well-defined desired counterfactual scenarios.

### 2.2 Data

#### 2.2.1 American Family Cohort

Our primary data source is the American Family Cohort (AFC), which is a research version of a Center for Medicare and Medicaid Services certified clinical registry and the largest primary care registry in the United States.^47^ AFC includes clinical, social, and demographic information for over 7 million individuals and 1300 practices, representing all 50 states. The dataset has a high representation of underserved (e.g., racial and ethnic minority, rural, and low-income) populations, and includes individuals insured through Medicare and Medicaid as well as privately. We used these primary care registry data to characterize stages of CKD progression, including among undiagnosed patients.

#### 2.2.2 Cohort definition

We defined a cohort of adult patients (i.e., age 18 years or older) for whom CKD progression can be observed in the AFC dataset between January 1, 2017 and December 31, 2017. Using standard codes,^48^ we extracted variables corresponding to age, binary recorded sex, serum creatinine measurements, diagnoses of CKD, diabetes, hypertension, and acute events known to impact creatinine levels (e.g., acute kidney injury, volume depletion, critical illness). Exclusion criteria were applied to remove those observed for less than a year after the inclusion date and those missing binary sex information. Extreme creatinine measurements above 73.8 and below 0 likely corresponded to other tests and were removed.^49^ Creatinine measurements captured within 30 days of acute events were also excluded, as they may not have been indicative of overall kidney health.^48^ We used the first available creatinine measurement to calculate eGFR values and subsequently classified individuals into CKD stages (eGFR ≥ 90: stage 1, 60-89: stage 2, 45-59: stage 3a, 30-44: stage 3b, 15-29: stage 4, ≤ 15: stage 5).^13^ Given the underutilization of urine tests necessary for establishing albuminuria status, our analysis depends solely on eGFR-defined staging. eGFR values below 5 were removed as they were unlikely to have been captured in a clinic. Additionally, we extracted census tracts corresponding to patient home addresses as well as recorded race and ethnicity.

#### 2.2.3 Social drivers of health

In addition to the AFC dataset, we considered two census tract-level indices of social deprivation and vulnerability: the Index of Concentration at the Extremes (ICE) and the Social Deprivation Index (SDI).^50,51^ These indices were generated using the 2020 American Community Survey data^52^ and assigned to individual patients based on census tracts. Individuals missing census tract information were excluded from the calculation of ICE and SDI calibration targets. The indices were selected to capture relationships between social factors known to impact the progression of CKD, based on prior literature.^11,25–27,53^ The ICE is a metric expressing concentrated extremes of both privilege and deprivation. Three types of ICE are available: income inequality, racial composition, or combined income and race. We used the latter, which jointly measures economic and racial segregation. For a given geographic area and population, it compares the fraction of non-Hispanic whites who are above the 80th percentile of income nationally with the fraction of non-white minorities whose income is below the 20th percentile. The SDI was developed to identify areas with unmet health care access needs for additional resource allocation. It is based on data regarding education, employment, family composition, housing quality, income, and transportation. We mapped the values of both indices into three quantiles based on their distribution in the AFC dataset.

### 2.3 Model

There are two primary simulation modeling approaches for CKD.^54,55^ The first creates discrete-time transitions through CKD disease states, defined by transition probabilities or risk equations^56,57^. The second considers continuous, linear eGFR decline, with decline rates typically sampled from predefined distributions.^58,59^ Both approaches allow for modeling changes in progression associated with time-dependent changes to diagnosed comorbidities, CKD diagnosis status, and interventions, such as particular treatments. However, because CKD disease states are defined by eGFR values in clinical practice, directly modeling eGFR decline is, in principle, clinically better motivated than discrete stage modeling.

We developed a continuous-time microsimulation model of eGFR decline to simulate a hypothetical cohort representing the AFC cohort, based on past studies, and data on social drivers. Model parameters were calibrated to reflect the CKD stage distributions in the AFC cohort conditional on sex, diabetes, hypertension, ICE quantiles, or SDI quantiles. The model allows for changes in progression associated with time-dependent changes to diagnosed comorbidities, CKD diagnosis status, and interventions, such as particular treatments. This process is represented by the conceptual flowchart in Figure 2. The model simulates individual eGFR trajectories over time, from initiation age of 30 until death. eGFR decline is accelerated by hypertension, diabetes and reaching CKD stage 3a. It can be delayed by interventions, which are applied according to a patient’s eGFR level, as measured by a particular eGFR formula.

**Figure 2:**
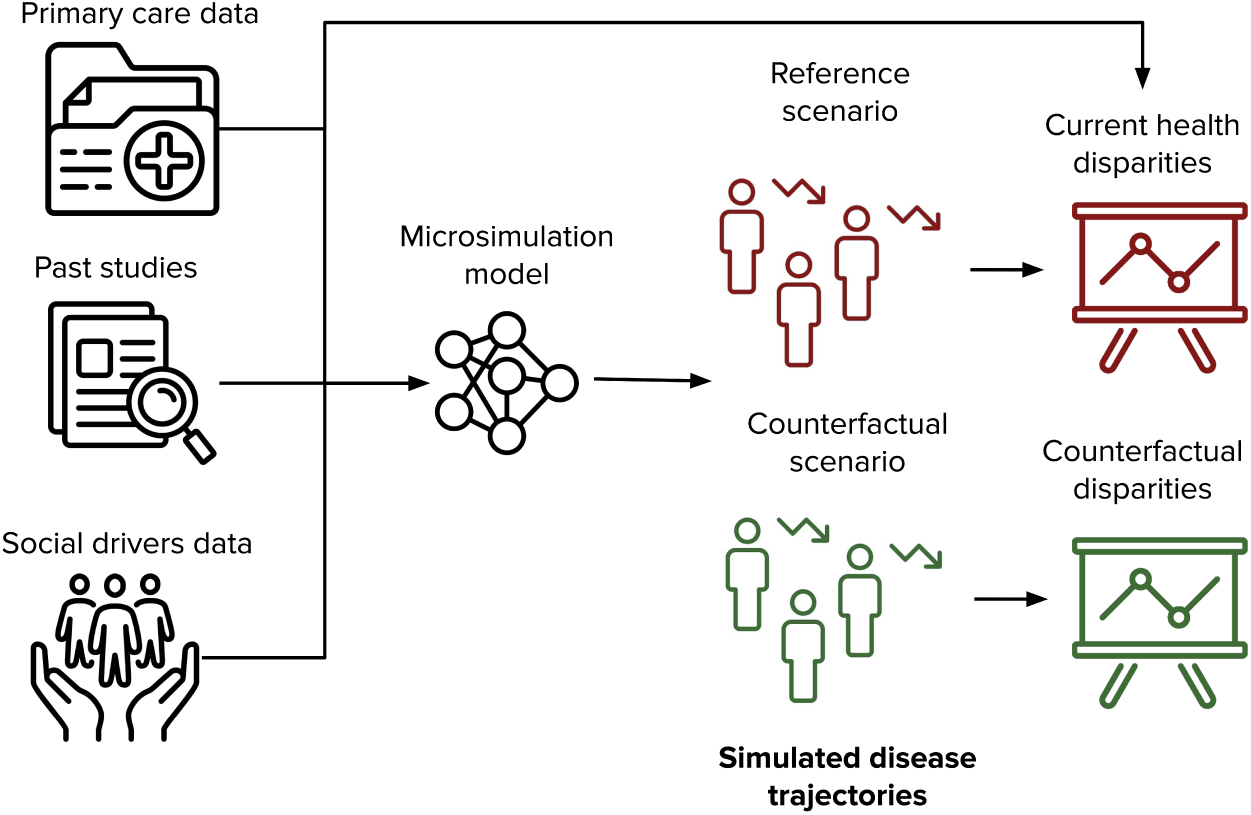
Conceptual flowchart representing the microsimulation model construction and the process of data simulation.

The model was then used to simulate the cohort under two scenarios: 1) reference, which corresponds to the setting under which the AFC data were collected when the race-adjusted eGFR formula would have been used, and 2) counterfactual, which reflects changes in time of treatment initiation following the switch to the 2021 CKD-EPI Creatinine-based eGFR equation (eGFR21) without race adjustment. While under the reference scenario, clinicians may have used one of several race-adjusted eGFR formulas. We assumed uniform use of the 2009 CKD-EPI Creatinine equation (eGFR09) for simplicity.^34^ All simulated individuals faced mortality risk specific to their age, sex, diabetes status, and eGFR level. The eGFR in the model corresponds to eGFR21, following current recommendations,^39,60^ and allows for ease of interpretation of model outputs by practitioners. The eGFR equations are included below. Additional details about parameter sources and modeling assumptions are included in the Parameters Supplement.

The eGFR21^39^ and eGFR09^34^ with serum creatinine (*S_cr_*) are:

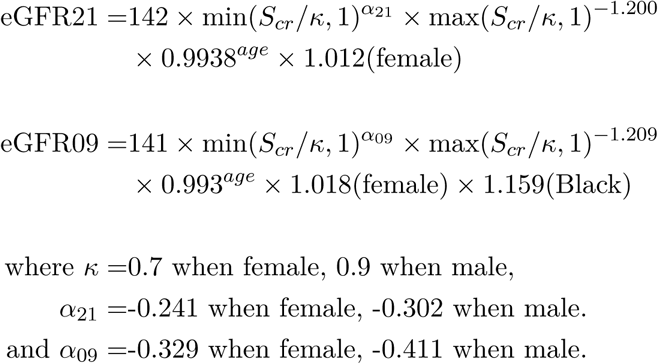

Trajectory simulation occurs in six steps, as shown in Figure 3. Rates of eGFR decline are conditional on individual-level covariates: progression to moderate or advanced CKD (stage 3a or above), incidence of diabetes and hypertension (see Table S3), and treatment status. Prior mean values of the decline rates were derived from previous analyses using NHANES data,^58,59,61^ and assumed the absence of albuminuria. Ages of diabetes of hypertension incidence were modeled using piecewise exponential frailty models, based on national incidence statistics grouped by age.^62,63^ The onset of hypertension additionally depended on sex. It was assumed that the timing of onset for both conditions was independent of one another.

**Figure 3:**
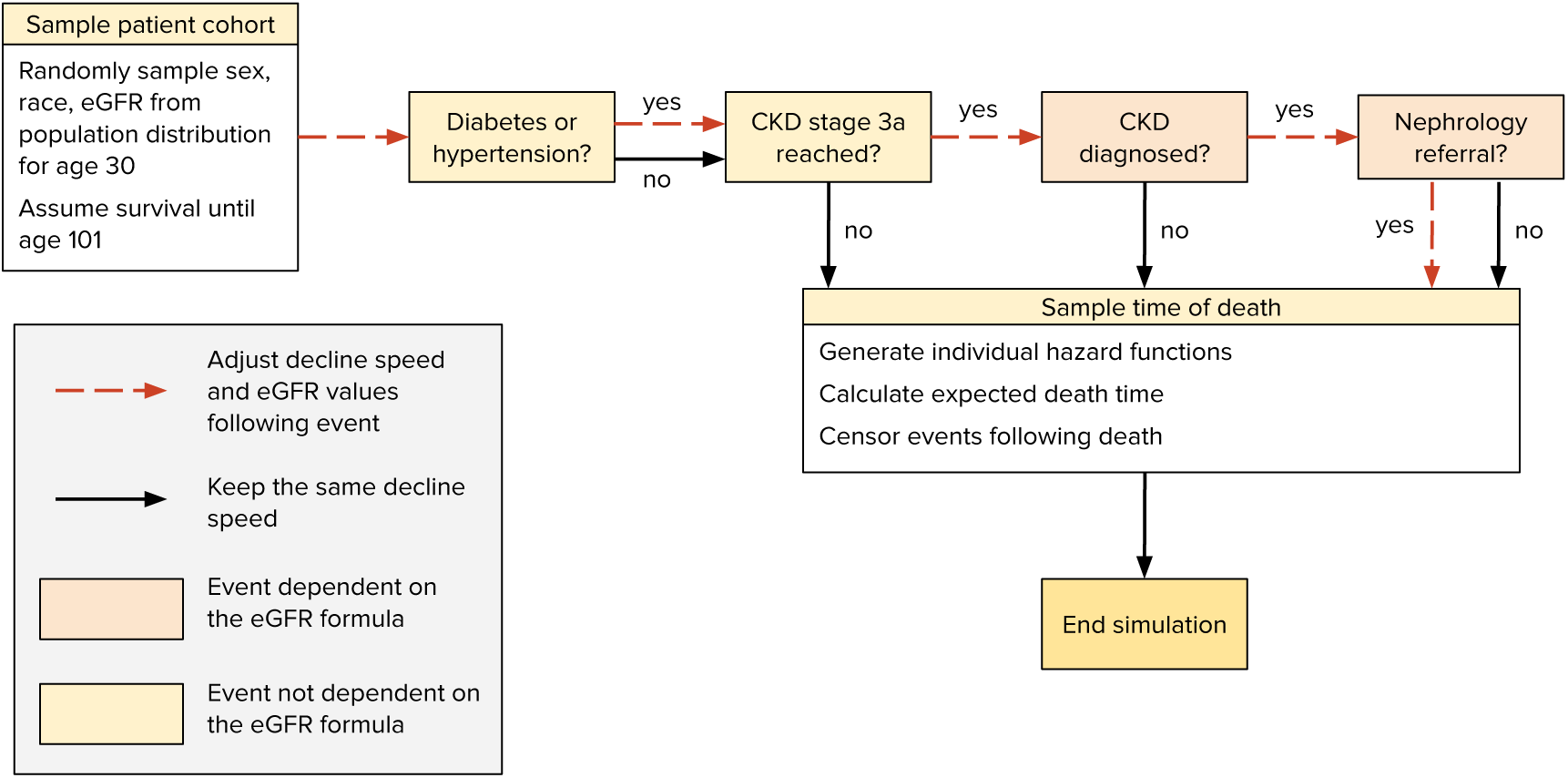
eGFR trajectory construction flowchart

We considered two interventions following a CKD diagnosis: enhanced comorbidity management and nephrology management. The model assumed that interventions can only be assigned starting at CKD stage 3a, with assignment probabilities increasing in more advanced stages, and that each individual assigned an intervention experienced the same reduction in the eGFR progression rate (Table S4). Interventions were applied the moment a patient’s eGFR crossed into a new stage and immediately resulted in reducing the speed of eGFR decline. Expected age of death was calculated from a piece-wise exponential hazard function obtained from age- and sex-specific life tables in 2019.^64^ These values were additionally adjusted with eGFR- and diabetes-specific hazard ratios.^65^ Further details of the model are included in the Model Supplement. We additionally considered the sensitivity of model outputs to changes in intervention frequency and effectiveness through sensitivity analysis, described in detail in the Sensitivity Analysis Supplement.

### 2.4 Calibration

Rates of eGFR decline conditional on diabetes, hypertension, and CKD stage could not be directly estimated from the data. To obtain them, we instead used a Bayesian calibration procedure using calibration targets derived from the AFC dataset, as illustrated in Figure 4. The targets reflect age-specific distributions of CKD stages by sex, diabetes, hypertension status, ICE quantiles, and SDI quantiles (Parameters Supplement).

**Figure 4:**
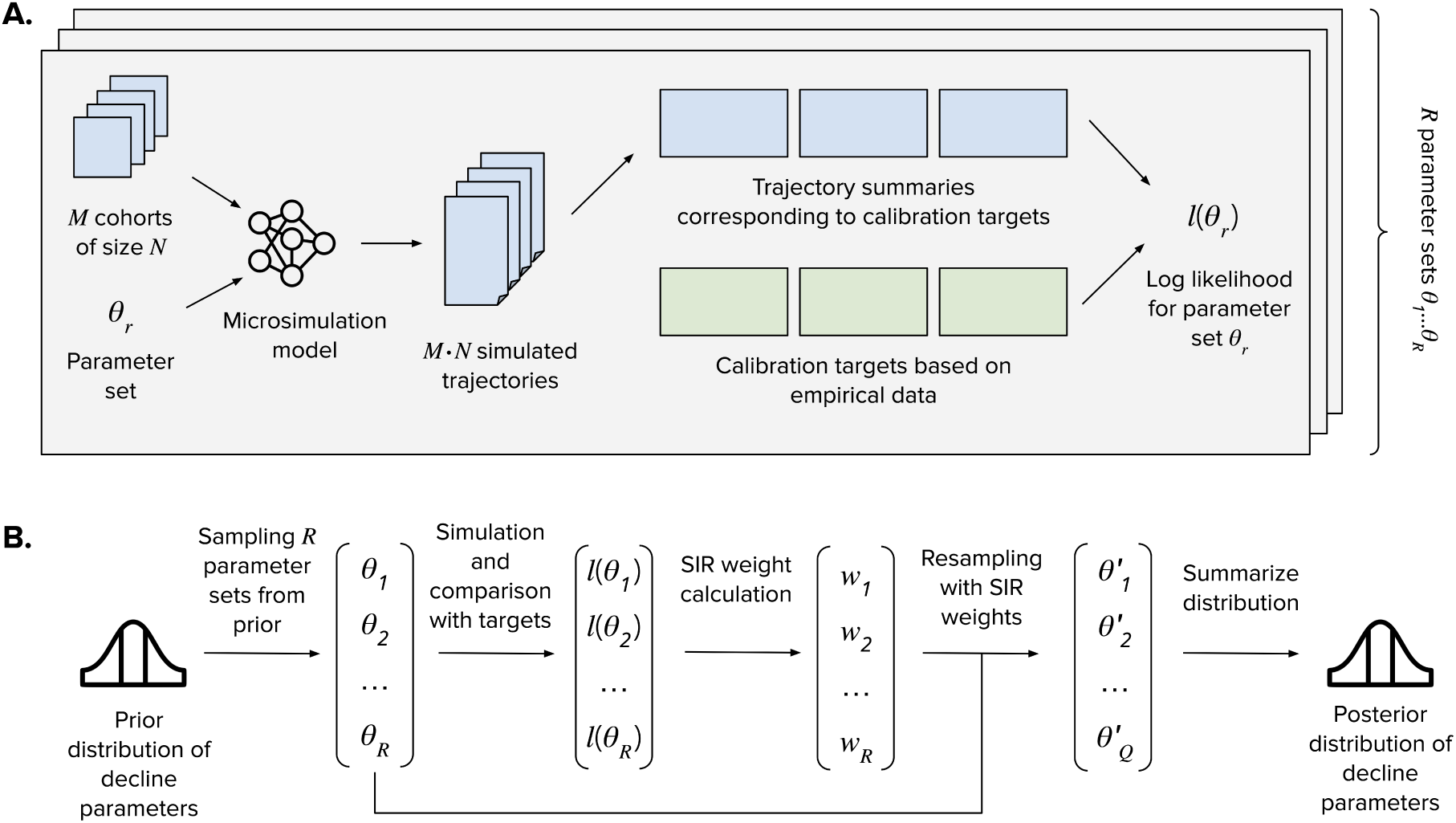
Calibration procedure. **A.** A parameter-set level log likelihood calculated by simulat-ing disease trajectories for *M* · *N* individuals across *M* sampled cohorts comparing their summaries to calibration targets using multinomial loss. **B.** Posterior of decline parameters calculated by sampling *R* parameter sets from the prior defined in Table S3, calculating parameter-set level log likelihoods following simulation, and using the sample importance resampling (SIR) procedure to weigh the *R* parameters based on their log likelihoods to obtain a posterior.

For all calibrated parameters, we defined truncated univariate normal prior distributions to exclude eGFR slopes indicating improvement over time, based on existing evidence, theory, and plausibility (Table S3). We applied a standard deviation corresponding to the coefficient of variation of 0.308 for sampling parameters. This coefficient corresponds to a standard deviation of 0.20 in the rate of progression among healthy individuals and captures the range of yearly rates of progression among healthy individuals reported in past literature.^58^ For combinations of covariates not previously reported^58^ (e.g., co-occurrence of diabetes and hypertension) we used the higher mean prior values corresponding to either one of the conditions occurring, and applied a higher coefficient of variation (0.461) to indicate a lower level of confidence in the priors. We further adjusted truncated normal priors based on regression analysis to achieve coverage of calibration targets.

We sampled *R* = 100, 000 parameter sets {*θ*_1_*, …θ_R_*} from the prior distributions using a Latin hypercube sampling design.^68^ To ensure that rates of decline increased with higher comorbidity burden and decreased with treatment, we used rejection sampling to sub-select parameter sets that followed that requirement. Cohorts of size *N* = 10, 000 were sampled, each composed of 50% men and 50% Black individuals, with *M* = 100 cohorts. We ran *R* · *M* experiments, generating sets of trajectories for each unique parameter set-cohort combination. Resulting trajectories were aggregated and compared against AFC calibration targets using a log-likelihood function comprising a sum of multinomial log-likelihoods, defined in the Model Supplement.

Model input parameter uncertainty for all outcome measures was accounted for by randomly sampling from the joint posterior distribution obtained from Bayesian calibration using the sample importance resampling algorithm.^69,70^ The posterior distribution was represented by a subset of sampled parameter sets with importance weights. We used 1000 parameter sets sampled from the posterior distribution to generate all primary outcomes for all scenarios and policies with 95% posterior model-prediction intervals for each outcome from the 2.5th and 97.5th percentiles of the projected values. Once the posterior distribution was identified, we recalculated eGFR trajectories for all *M* cohorts in the counterfactual scenario corresponding to the posterior, and compared them with regards to life expectancy, distribution of CKD stages across ages, and eGFR value at intervention, stratified by sex, race, ICE, and SDI.

## 3 Results

### 3.1 Data summaries

We extracted a cohort of 733,337 individuals from the AFC dataset, described in Table 1. A cohort extraction flowchart also appears in the Figures and Tables Supplement. The cohort has a mean age of 60 and is 44% male. At inclusion, 8% of individuals had a CKD diagnosis code. This is lower than the national age-adjusted prevalence of 21%, but consistent with a high degree of underdiagnosis of CKD.^12^ Additionally, 25% of individuals had a diabetes diagnosis and 60% had a hypertension diagnosis, similar to national prevalence values. Our cohort had 88% of individuals with an eGFR value at or above 60, corresponding to no CKD or stages 1 and 2.^62,63^ Only 6% of our cohort was Black or African American, with 79% white individuals. Of note, 12% of the cohort were missing race, 27% were missing ethnicity information, and 15% had missing census tract information. For the social indices, ICE and SDI, we observed a health gradient, where indices indicating higher levels of deprivation were associated with higher prevalence of diabetes, hypertension, and CKD. For instance, the prevalence of diabetes ranged from 19% to 31% in the least and most deprived ICE quantile, respectively.

**Table 1:**
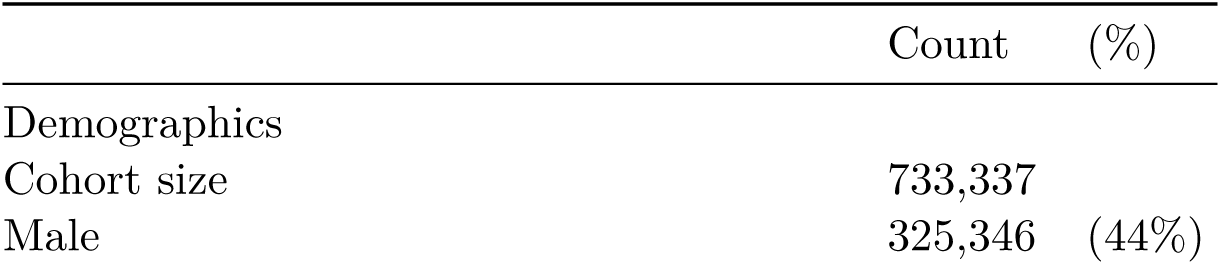
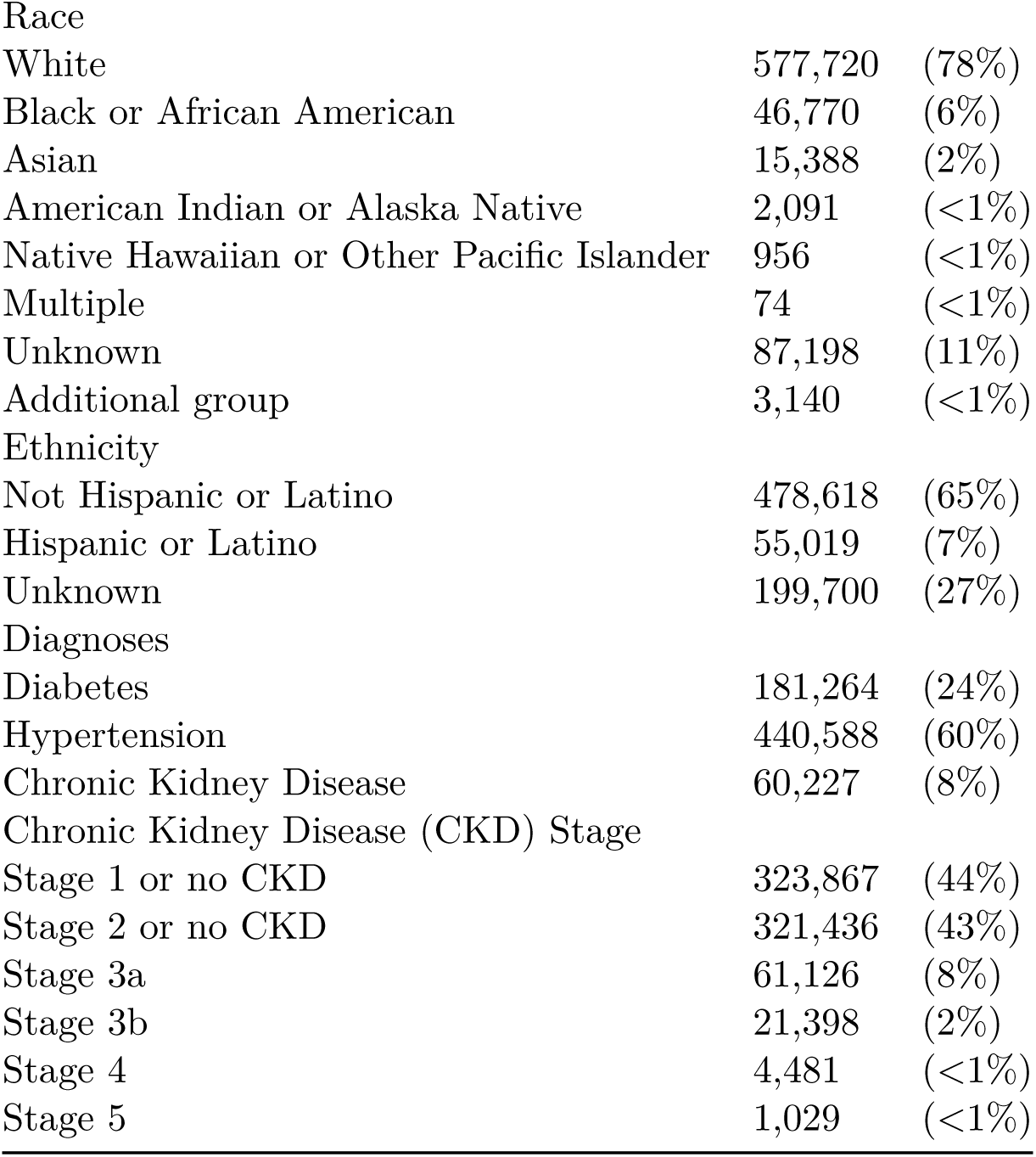
American Family Cohort data summary

|  | Count | (%) |
| --- | --- | --- |
| Demographics |  |  |
| Cohort size | 733,337 |  |
| Male | 325,346 | (44%) |
| Race |  |  |
| White | 577,720 | (78%) |
| Black or African American | 46,770 | (6%) |
| Asian | 15,388 | (2%) |
| American Indian or Alaska Native | 2,091 | (<1%) |
| Native Hawaiian or Other Pacific Islander | 956 | (<1%) |
| Multiple | 74 | (<1%) |
| Unknown | 87,198 | (11%) |
| Additional group | 3,140 | (<1%) |
| Ethnicity |  |  |
| Not Hispanic or Latino | 478,618 | (65%) |
| Hispanic or Latino | 55,019 | (7%) |
| Unknown | 199,700 | (27%) |
| Diagnoses |  |  |
| Diabetes | 181,264 | (24%) |
| Hypertension | 440,588 | (60%) |
| Chronic Kidney Disease | 60,227 | (8%) |
| Chronic Kidney Disease (CKD) Stage |  |  |
| Stage 1 or no CKD | 323,867 | (44%) |
| Stage 2 or no CKD | 321,436 | (43%) |
| Stage 3a | 61,126 | (8%) |
| Stage 3b | 21,398 | (2%) |
| Stage 4 | 4,481 | (<1%) |
| Stage 5 | 1,029 | (<1%) |

### 3.2 Model calibration

Our calibration procedure generated a single best-fitting parameter set, which we refer to as the mean posterior. The inclusion of ICE and SDI calibration targets did not impact the value of the mean posterior. Figure 5 shows the value of the mean posterior compared to the mean prior slope parameters, as well as the distribution of sampled parameters. Mean baseline rate of decline among healthy individuals was 0.68 mL/min/1.73m^2^, 5% higher than that in the prior, and increased by 13% after reaching CKD stage 3a (compared to no change in the prior). Decline prior to CKD stage 3a was elevated 1% by comorbid diabetes, 15% by hypertension, and 159% by a combination of both (compared to 69%, 11% and 69% increase in the prior). Decline after reaching CKD stage 3a was elevated 152% by comorbid diabetes, 24% by hypertension, and 163% by a combination of both (compared to 331%, 115%, and 331% increase in the prior).

**Figure 5:**
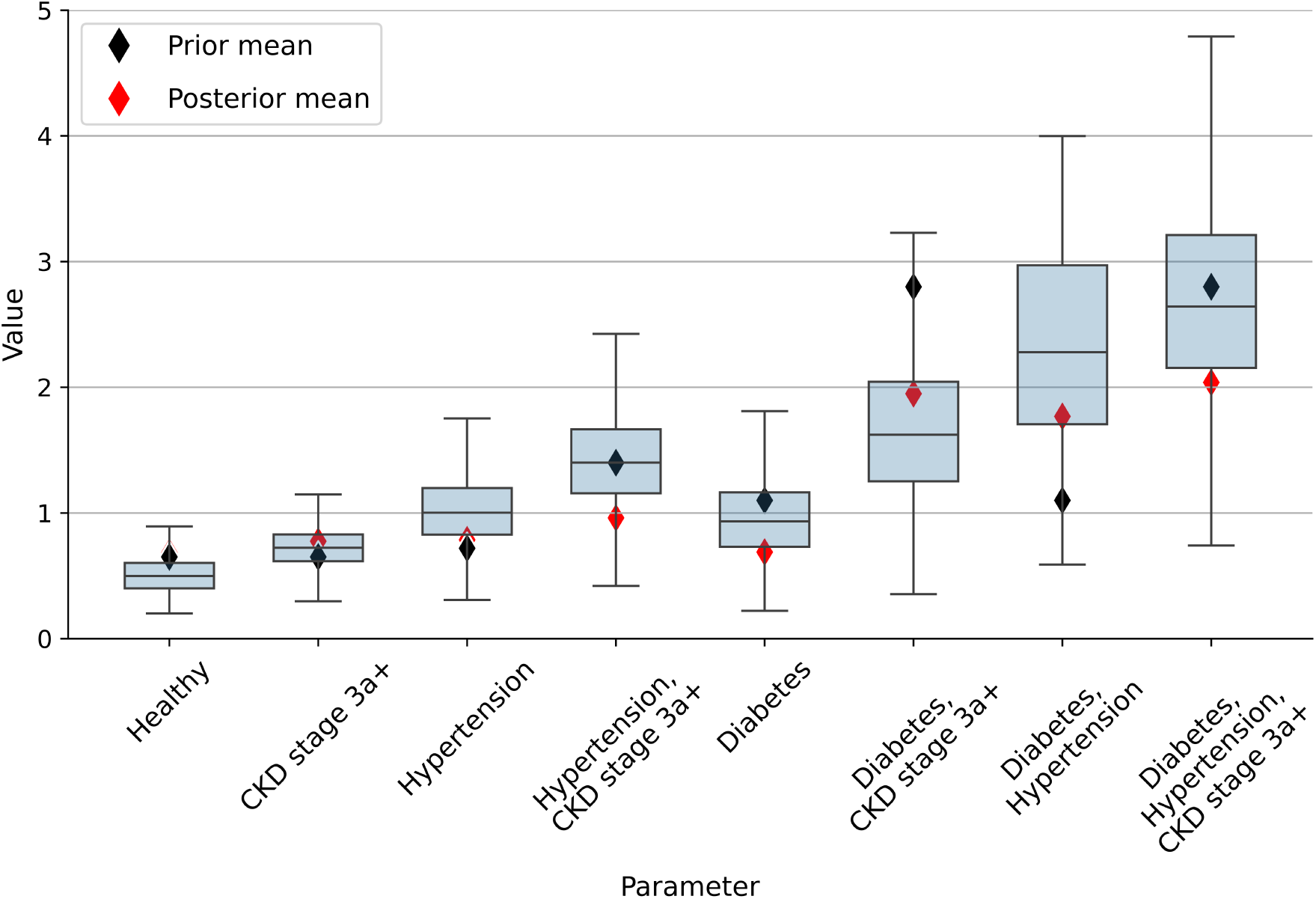
The distribution of sampled parameters (blue), with mean prior^58^ (in black) and posterior (in red) values marked. Outlier values are not shown. Healthy corresponds to individuals in chronic kidney disease (CKD) stages 1 and 2 or without CKD, who also do not have diabetes or hypertension.

We examined the distribution of individuals across CKD stages and ages stratified by sex (Figure S6), diabetes (Figure S7), and hypertension (Figure S8) for both simulation scenarios, comparing the prevalence observed in our AFC cohort corresponding to calibration targets. Both simulation scenarios generated highly similar, overlapping results. Prevalence was closely matched to that in the AFC cohort in CKD stages 1 and 2 for the sex strata, as well as for individuals with diabetes or hypertension, and less closely matched for those without diabetes or hypertension. Results were more imprecise at later ages and later stages, where group sizes were small. In particular, lower prevalence at later ages in stages 3a and 3b and higher in stages 4 and 5 in our simulations.

### 3.3 Simulation results

We compared mean life expectancy at age 30 under our two simulated scenarios, separately considering groups stratified by sex, race and CKD status, and included the results in Table 2. Under the model assumptions, Black individuals would be expected to survive longer, and non-Black individuals shorter, under the counterfactual scenario compared to the reference. The magnitude of differences was more pronounced for non-Black individuals although differences did not surpass two months for any group. The sensitivity analysis revealed that even among those with CKD, under immediate and uniform diagnosis starting at stage 3a and increased treatment effectiveness, differences in life expectancy would not exceed 4.2 months.

**Table 2:** Mean additional life expectancy (in months) under the counterfactual scenario compared to the reference scenario, for individuals in the simulated population.

| Maximum CKD stage | Black Female | Black Male | Non-Black Female | Non-Black Male |
| --- | --- | --- | --- | --- |
| Any stage | 0.15 | 0.12 | -0.63 | -0.58 |
| Stage 3a or later | 0.45 | 0.44 | -1.50 | -1.64 |

In our main results, we compared the earliest times at which simulated individuals would qualify for a diagnosis at each CKD stage for the two scenarios (Figure 6).Under the counterfactual scenario with eGFR21, Black individuals would be eligible for diagnosis earlier, and non-Black individuals later, compared to the reference eGFR09 scenario. The difference was largest for earlier stages and smaller at each consecutive CKD stage. For example, under the counterfactual, the earliest diagnosis into stage 2 would on average be 9.6 and 9.1 years earlier for Black women and Black men, but 4.4 and 4.8 years later for non-Black women and non-Black men. However, the earliest diagnosis into stage 5 would, on average, be 0.7 and 0.6 years earlier for Black women and Black men, but 1.1 years later for non-Black women and non-Black men. We also compared the difference in eGFR values that would qualify individuals into particular stages under the two scenarios. Under the counterfactual scenario, Black individuals would be eligible for diagnosis at higher values of eGFR with non-Black individuals at lower values than under the reference. Similar to the difference in diagnosis times, the differences in eGFR values between the two scenarios decreased at each consecutive stage.

**Figure 6:**
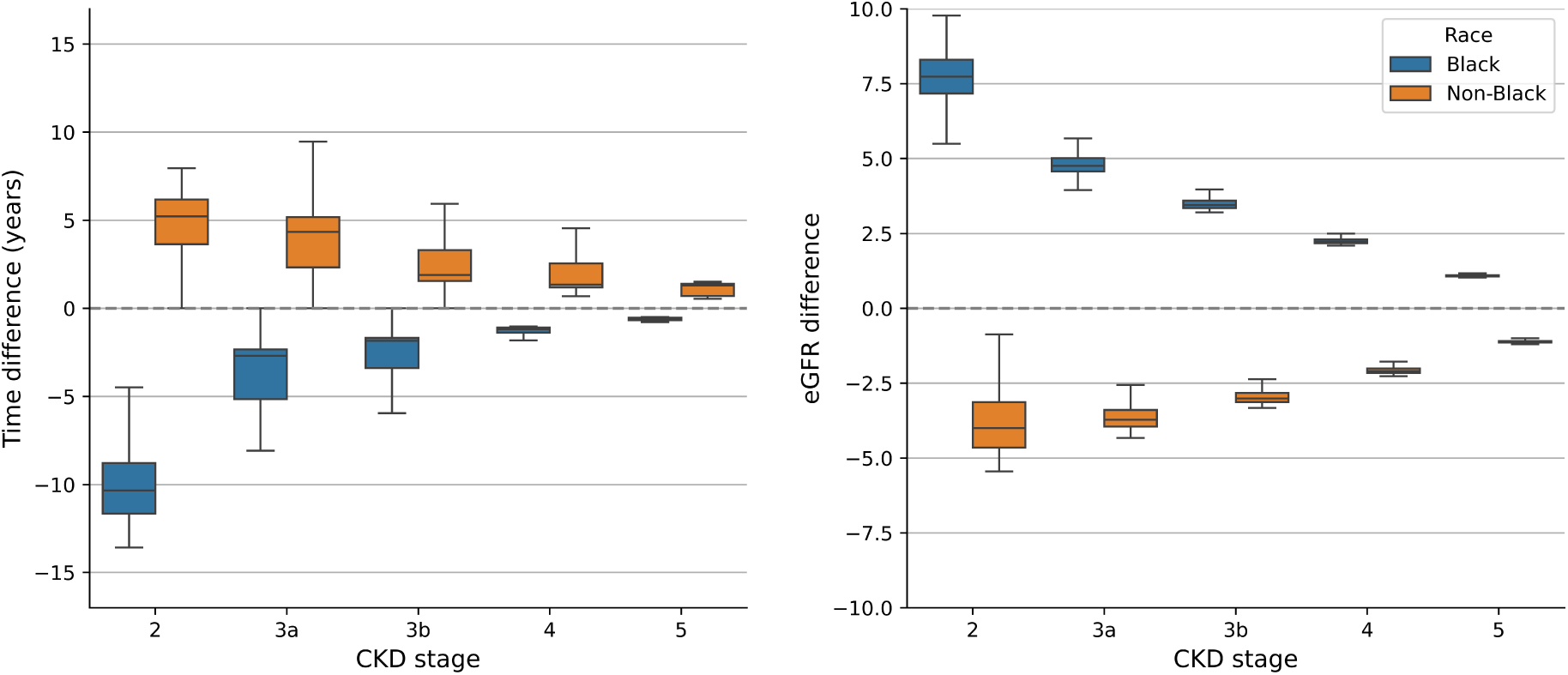
Difference in time (years) and eGFR (mL/min/1.73m^2^) value at the earliest possible diagnosis to a given chronic kidney disease (CKD) stage under eGFR21−eGFR09. Negative values indicate earlier diagnosis (left) or lower value of eGFR during diagnosis (right) under eGFR21 compared to eGFR09. Outliers not shown.

## 4 Discussion

We developed a framework for mitigating societal bias in data, creating new values under a pre-specified data generating process in a first-of-its-kind microsimulation-based data transformation method. This involved generating a microsimulation model of CKD progression based on eGFR decline over time, calibrated to a cohort of patients in a large primary care dataset. Our model was able to reproduce stage distributions observed in the cohort, which reflected patterns of CKD progression and care informed by the 2009 CKD-EPI equation.

The model allows for generating counterfactual eGFR trajectories that reflect the use of the 2021 CKD-EPI equation through adjusting the timing of interventions based on the counterfactual eGFR levels. The trajectories simulated under the counterfactual scenario reflected earlier diagnoses for Black patients and later diagnoses for non-Black patients than those observed in the data. However, these changes led to differences in life expectancy not exceeding 2 months among those with CKD. While these results were sensitive to assumptions on the rate of diagnosis and intervention effectiveness, the simulated effect among those with CKD did not exceed 4.2 months even under assumptions of universal diagnosis and treatment initiated at stage 3a.

The simulated data could be used directly as inputs into predictive algorithms for a number of outcomes, including timing of CKD incidence, CKD progression speed, time of diagnosis initiation and nephrology referral, time of reaching ESRD,^71^ and mortality. This goes beyond reclassification and can include effects of treatment in the short and long term. These outcomes can be easily defined due to the continuous disease trajectories. The framework also allows for a flexible adaptation to other counterfactual scenarios, such as changes in diagnosis frequency or regional differences in frequency of nephrology referrals. It can additionally be used to up-sample underrepresented populations, such as those residing in areas with lower access to nephrology care.^72,73^ Future work could explore realistic sub-sampling of measurement times to reflect a real-world practice of collecting discrete measurements, as well as adding patterns of missingness.

While our results suggest that the removal of the race adjustment from the eGFR equation is likely to lead to notable changes in diagnosis eligibility in earlier stages, those changes correspond to small changes to life expectancy. As such, the change of the eGFR equation by itself is unlikely to reduce the burden of CKD among Black Americans and reduce disparities in CKD outcomes in the United States. Our sensitivity analysis suggests that effects would remain modest (not exceeding 4.2 months of additional life expectancy) even under perfect guideline concordance regarding early diagnosis and treatment of CKD.

The differences in earliest possible diagnosis time in the two simulation scenarios followed the hypothesized direction from prior literature,^39^ with Black patients qualifying for diagnosis earlier and non-Black patients later than in the reference scenario. However, the actual time of diagnosis is likely to lag behind the earliest possible diagnosis time because it depends on the clinician’s decision to initiate diagnosis and requires two blood tests separated by at least 90 days to establish chronicity.^14^ The difference is much higher in earlier CKD stages, where diagnoses are less frequent. In stage 2, where differences were largest, additional urine testing is needed to establish a diagnosis. The observed effect on the timing of diagnoses may therefore be smaller than reported and modified by factors related to the health system and health access. This is also suggested by a recent study at Stanford Health Care that demonstrated that adopting the new eGFR equation without race adjustment did not impact rates of nephrology referrals and visits after two years.^74^ Our results additionally point to potential adverse consequences of the change in the eGFR equation among non-Black patients, who could experience delayed care and slightly elevated mortality compared to the 2009 criteria. This is consistent with the evidence that the formula change on average leads to eGFR overestimation for non-Black patients, and eGFR underestimation for Black patients.^24,75,76^

Rates of progression identified through the Bayesian calibration procedure differed from those previously derived from NHANES data.^58,59^ In particular, rates of progression following CKD stage 3a, while higher than those in earlier stages, did not increase as notably in our model as in NHANES data. This could potentially reflect a higher quality of CKD and comorbidity management among the AFC population compared to the national sample. The rates did not differ across area-level social deprivation indices, which might be explained by similarity between index-specific calibration targets.

Our work has several limitations. The AFC dataset included short observation periods for individuals, high variability in the frequency of creatinine observations, and data coding errors common in electronic health data. Given the limited data on albuminuria available in the AFC dataset, we also did not include albuminuria status in our model, as CKD models often do.^54^ Additionally, our choice of an eGFR-based CKD progression model, considered better clinically motivated than discrete stage modeling,^55^ made it possible for us to identify the timing of eGFR-based interventions and their counterfactuals more directly. However, it prevented us from using the complete set of intermittently observed data in the AFC dataset to inform our model, in ways that a stage-based model may have allowed. We assumed a uniform stage-conditional probability of diagnosis and nephrology treatment although those differ across states, race and ethnicity, age, socioeconomic status, and insurance status.^72,73^ Future analyses could consider differences in rates of diabetes and hypertension incidence as well as CKD diagnosis and nephrology referrals across social deprivation index quantiles. Further, we assumed that interventions would be triggered immediately after crossing an eGFR threshold value. In practice, interventions would typically be initiated with some delay, based on the timing of clinician visits, would likely not be effective immediately, and would be subject to discontinuation by some patients. The set of interventions available for CKD patients is vast and their matching to patient profiles is complex. Our consideration of two interventions limited the range of effects observed. We also assumed treatment can incur only benefits, so potential harms resulting from over-treatment are not reflected in our results. Prior literature reports a wide range of treatment effectiveness values, and our assumption of uniform effectiveness may have impacted our results. Finally, we note that formulas without race adjustment that include cystatin C have reported smaller discrepancies between estimated and measured GFR for both groups than equations considered here.^39^ Such equations have not seen a broad uptake due to cost-effectiveness concerns.

Our data transformation framework demonstrates how the explicit representation of the data generation process can help anticipate the effect that policy changes have on clinical data distributions. By simulating continuous disease trajectories, and explicitly modeling clinical decisions and their effectiveness, our framework can be used to generate a range of counterfactual values.

## Acknowledgements

We thank Malcolm Barrett, Oana Enache, and Sara Khor for their valuable insights and contributions to code review.

The following acknowledgment text is included as described by the Stanford Center for Population Health Sciences Data Core (phsdocs.stanford.edu/v1.0/need-help/citing-phs-data-core): “Data for this project were accessed using the Stanford Center for Population Health Sciences Data Core. The PHS Data Core is supported by a National Institutes of Health National Center for Advancing Translational Science Clinical and Translational Science Award (UL1TR003142) and from Internal Stanford funding. The content is solely the responsibility of the authors and does not necessarily represent the official views of the NIH.”

## Statements and Disclosures

### Ethical considerations

This study obtained approval from the Institutional Review Board at Stanford University.

### Consent to participate

Not applicable

### Consent for publication

Not applicable

### Data availability

The Python code and summary data to reproduce our results are available at github.com/StanfordHPDS/data_transformation. All analyses described in the manuscript can be reproduced, with the exception of the generation of data summaries and calibration targets, which require access to the AFC dataset. The AFC dataset contains protected health information and cannot be shared publicly.

## S1: Model Supplement

### Sampling time to diabetes and hypertension

We modeled time to comorbidities (i.e., diabetes and hypertension, separately) using piecewise exponential frailty models, based on incidence statistics grouped by age.^62,63^ Let *F^c^* be the cumulative distribution function (CDF) of developing a comorbidity *C*:

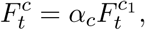

where *F*^*c*1^_*t*_ is the CDF of those that eventually develop the comorbidity, and *α_c_* is the proportion of individuals who eventually develop the comorbidity in their lifetime. For each *i*th simulated individual, we sampled whether they will eventually develop the comorbidity *C* following a Bernoulli distribution:

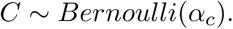

For those who would eventually develop the comorbidity (*C* = 1), we sampled the age of comorbidity onset:

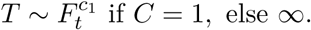

Time to diabetes was not conditional on any additional covariates. We estimated the CDF of developing diabetes, *F^d^_t_* = *α_d1_F^d^_t_*, based on CDC yearly incidence statistics (Table S1),^62^ by treating yearly incidences as hazards *h^d^_t_* that apply across age ranges:

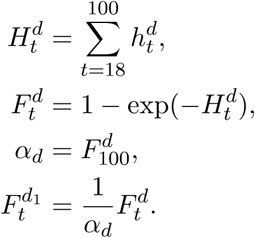

We assumed that *α_d_* corresponds to the CDF at age 100. Time to hypertension was conditional on sex only (Table S1).^63^ We did not consider the joint probability of developing diabetes and hypertension, given availability of data sources.

### Time of death calculation

Expected age of death *T* was calculated from a piece-wise exponential hazard function obtained from age- and sex-specific life tables in 2019.^64,77^ For each *i*th individual, their corresponding mortality hazard function *h*(*t*) was adjusted to reflect the impact of individual covariates *X*(*t*), CKD status and diabetes, at time *t* on background hazard function *h_b_*(*t*), assuming proportional hazards. Hazard ratios HR(*X*(*t*)) applied to all-cause mortality, irrespective of albuminuria status overall,^65^ and were used once one’s eGFR value reaches 60 (Table S2). This resulted in a unique hazard function *h*(*t*) for each individual based on their eGFR trajectory, which was then used to obtain their expected age of death, conditional on survival until 30, using life table equations detailed below:

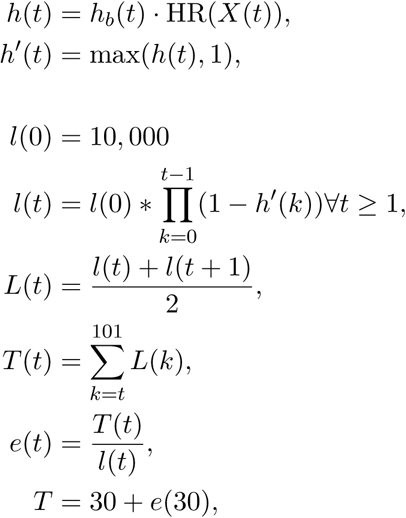

where *l*(0) is the size of the hypothetical population at the time of birth, *l*(*t*) is the number of individuals alive at the beginning of the age interval, *L*(*t*) is the number of person-years lived by *l*(*t*) individuals during the age interval, *T* (*t*) is total person-years accrued by cohort beyond age *t*, and *e*(*t*) is the average remaining lifetime for an individual alive at the beginning of age interval *t*.

### Simulating counterfactual outcomes

Reference and counterfactual scenarios for an individual differed primarily by the time at which progression-delaying interventions are applied. Hence, they differed in the portions of trajectories following interventions. For those receiving interventions at a specific stage, a change in the speed of eGFR decline was applied when the eGFR value reached a specific cutoff corresponding to that stage. That threshold was given by the eGFR09 equation under the reference scenario and by the eGFR21 equation under the counterfactual scenario. Counterfactual trajectories additionally differed from reference trajectories by time of death. The mortality hazard function for the counterfactual was adjusted with hazard ratios corresponding to eGFR values following an intervention, resulting in a divergence of survival functions. Using common random numbers, reference and counterfactual death times were sampled jointly from each individual’s survival functions. For those not assigned interventions, either because they never progressed into a more advanced CKD stage or because they progressed but were not diagnosed and treated, the two scenarios resulted in identical trajectories.

### Calibration

#### Sampling parameters from a prior

We calibrated the model with respect to *L* = 8 parameters, which described annual rates of eGFR decline conditional on 3 binary variables: diabetes, hypertension, and moderate-or-advanced CKD. The parameter sets came from a vector of truncated univariate normal prior distributions defined in Section 2.4 and Table S3:

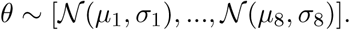

We sampled *R* parameter sets [*θ*_1_, *. . . θ_R_*] from the prior before the start of the experiment and then saved them.

#### Calibration targets

Calibration targets corresponded to the distribution of individuals across CKD stages, and are defined in the Parameters Supplement. Each of the *S* = 5 calibration targets corresponded to *A_s_* multinomial distributions. Each multinomial distribution *a* was defined by *N_s,a_* trials and *K* = 6 mutually exclusive events, where *N_s,a_* was the size of the cohort in stratum *a* for calibration target *s* (defined by covariates such as age) and *K* is the number of stages. For example, the first calibration target consisted of *A*_1_ = 14 individual multinomial distributions, each corresponding to a distribution of individuals within a single age category and a single sex across six CKD stages. As such, for each calibration target *s* and stratum *a*, the number of individuals in each stage was represented from the data as **x_s_**,**_a_**, defined as:

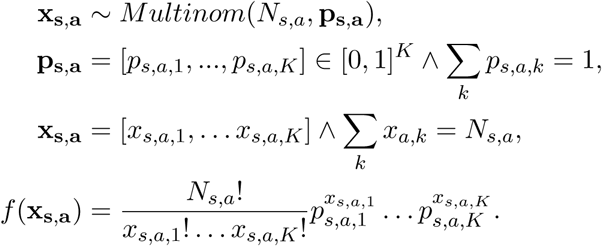

In our experiments, **x_s_**,**_a_** come directly from empirical data, and are defined in Table S5, Table S6, Table S7, Table S8 and Table S9. Parameters **p_s_**,**_a_** were generated from the simulation model as **p_s_**,**_a_** = **q_s_**,**_a_***/Q_s,a_*, where **q_s_**,**_a_** was the simulated number at each stage and **1**^⊺^**q_s_**,**_a_** = *Q_s,a_*.

#### Running an experiment matrix

We ran *R* · *M* experiments generating *M* · *N* trajectories for each parameter set *θ_r_*. Summary matrices *q_s,a,k_*(*θ_r_, m*) were calculated for each calibration target *s* by considering the distribution of stages and comorbidities every 10 years (at ages 35, . . . , 95) corresponding to 10-year age bins and selected binary categories. We aggregated matrices by taking an average count across all *M* cohorts, and calculated summary trials values *Q_s,a_*(*θ_r_*) as well as distribution frequencies p_s,a_(*θ_r_*) as defined below:

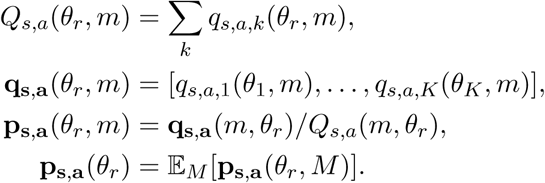

#### Coverage analysis

To assesed the coverage of calibration targets by experiment summaries, we calculated 95% Wald-type confidence intervals on the calibration targets, using the following formula:

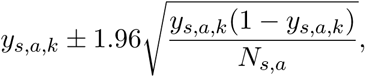

where *y_s,a,k_* = *x_s,a,k_/N_s,a_* with *x_s,a,k_* and *N_s,a_* coming directly from AFC cohort.

We calculated the 95% uncertainty bounds (using the 95% interquantile range) for the simulated outcomes, using the 2.5th and 97.5th quantile for each *p_a,k_* directly from its distribution across all parameter sets (*P_a,k,r_*).

#### Calculating the log-likelihood function

The log-likelihood *l_s,a_*(*θ_r_*|p*_s,a_*(*θ_r_*)) of the parameter set *θ_r_* generating x_s,a_(*θ_r_*) with respect to the calibration target *a* and a calibration strata *s* is defined below. We combined those log-likelihoods across strata and calibration targets with simple summation to generate *l*(*θ_r_*):

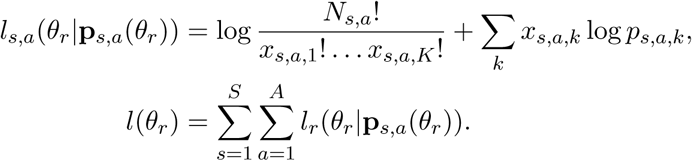

The log likelihood can become infinitely negative when any of the *p_k_* = 0. We removed those rows from calibration targets, generating multinomials with fewer categories for some strata.

#### Calculating the posterior with sample importance resampling

Once we calculated parameter set-level log-likelihoods *l*(*θ_r_*), we computed sampling importance weights *w_r_* using a softmax function. We then resampled with replacement *Q* = 1000 times from the discrete distribution [*θ*_1_, *…, θ_R_*] using sample importance resampling weights, obtaining a matrix *θ^′^* = [*θ^′^ , . . . θ^′^*]:

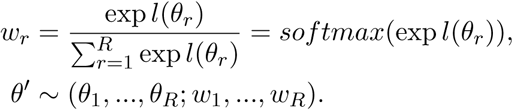

Because we assumed that each of the L parameters was an independent, normally distributed variable, the posterior distribution of decline parameters *θ^′^* could be calculated by separately computing means and standard deviations for each dimension *l* across the *Q* resampled parameter sets in *θ^′^*:

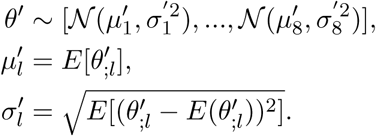

## S2: Parameters Supplement

The initial distribution of eGFR for individuals was set at N (108.8, 15.5), corresponding to the distribution in the AFC cohort among individuals aged 25-35. Onset of hypertension and diabetes was based on annual hazards of incidence, as shown in Table S1.^62,63^ For hypertension, we additionally assumed that incidence in ages 18-19 and 80-100 matches that in ages 20-29 and 70-79. Annual hazard rates for background mortality came from sex-specific 2019 life tables.^64^ Previously reported hazard ratios^65^ were used to additionally account for increased mortality associated with diabetes and eGFR level below 60 (Table S2). The ratios correspond to mean hazard ratios for all-cause mortality, irrespective of albuminuria status. Mortality adjustments were not applied above eGFR 60 given the lack of clear explanation for elevated mortality at higher eGFR levels.

**Table S1:** Annual rates of diabetes and hypertension incidence, based on published values^62,63^ and additional assumptions.

|  | Sex | Age | Annual rate |
| --- | --- | --- | --- |
| Diabetes | Any | 18-44 | 0.003 |
|  |  | 45-64 | 0.0101 |
|  |  | 65-100 | 0.0068 |
| Hypertension | Male | 18-29 | 0.0055 |
|  |  | 30-39 | 0.0166 |
|  |  | 40-49 | 0.0219 |
|  |  | 50-59 | 0.0236 |
|  |  | 60-69 | 0.028 |
|  |  | 70-100 | 0.0311 |
|  | Female | 18-29 | 0.002 |
|  |  | 30-39 | 0.0077 |
|  |  | 40-49 | 0.018 |
|  |  | 50-59 | 0.0249 |
|  |  | 60-69 | 0.0347 |
|  |  | 70-100 | 0.0428 |

Parameters for interventions came from a combination of past studies and assumptions. We assumed primary care management following CKD diagnosis starting in stage 3a. Diagnosis rates for stages 3a and 3b were from an analysis of records of patients meeting diagnostic criteria in large electronic health record databases.^78^ We do not consider differences in diagnosis rate by sex reported by the study. Diagnosis rates in stages 4 and 5 were from a smaller analysis^79^.

Frequencies of nephrology management in stages 3a, 3b and 4 were obtained from a retrospective multicenter study of stage 3 and 4 patients in Massachussetts.^73^ These values only consider diagnosed patients from a state with one of the highest rates of pre-ESRD nephrology care in the US.^72^ Frequency of management in stage 5 came from national analyses of the US Renal Data System, and corresponds to the fraction of patients who reported receiving some form of nephrology care prior to end stage renal disease requiring dialysis.^72,80^

**Table S2:** Mortality hazard ratios, based on eGFR and diabetes.^65^

| eGFR | Diabetes | No diabetes |
| --- | --- | --- |
| $\geq 60$ | 1 | 1 |
| 45-59 | 1.18 | 1.19 |
| 30-44 | 1.65 | 1.53 |
| 15-29 | 2.28 | 2.27 |
| 0-14 | 4.46 | 4.06 |

**Table S3:** Mean prior rates of annual eGFR decline [mL/min/1.73m ^2^/year] among groups defined by chronic kidney disease (CKD) stage and presence of comorbidities.^58^

| Comorbidities |  |  | Mean annual eGFR decline |
| --- | --- | --- | --- |
| Before CKD stage 3a | No hypertension | No diabetes | 0.65 |
|  |  | Diabetes | 1.10 |
|  | Hypertension | No diabetes | 0.72 |
|  |  | Diabetes | 1.10 |
| CKD stage 3a and later | No hypertension | No diabetes | 0.65 |
|  |  | Diabetes | 2.80 |
|  | Hypertension | No diabetes | 1.40 |
|  |  | Diabetes | 2.80 |

Precise estimates of the effectiveness of considered interventions were not available in literature. Average change in the rate of decline among CKD patients receiving primary care was approximately 77% (from 3.20 to 0.74ml/min/1.73m^2^/year).^81^ A small study reported that the initiation of nephrology care resulted in an average reduction in the rate of decline from -5.4 to -0.35 ml/min/1.73m^2^/year, corresponding to a 94% reduction.^19^ We adjusted these estimates, assuming a reduction of the speed of eGFR decline of 20% as a result of enhanced primary care management following CKD diagnosis and of 60% following the initiation of nephrology care.

**Table S4:**
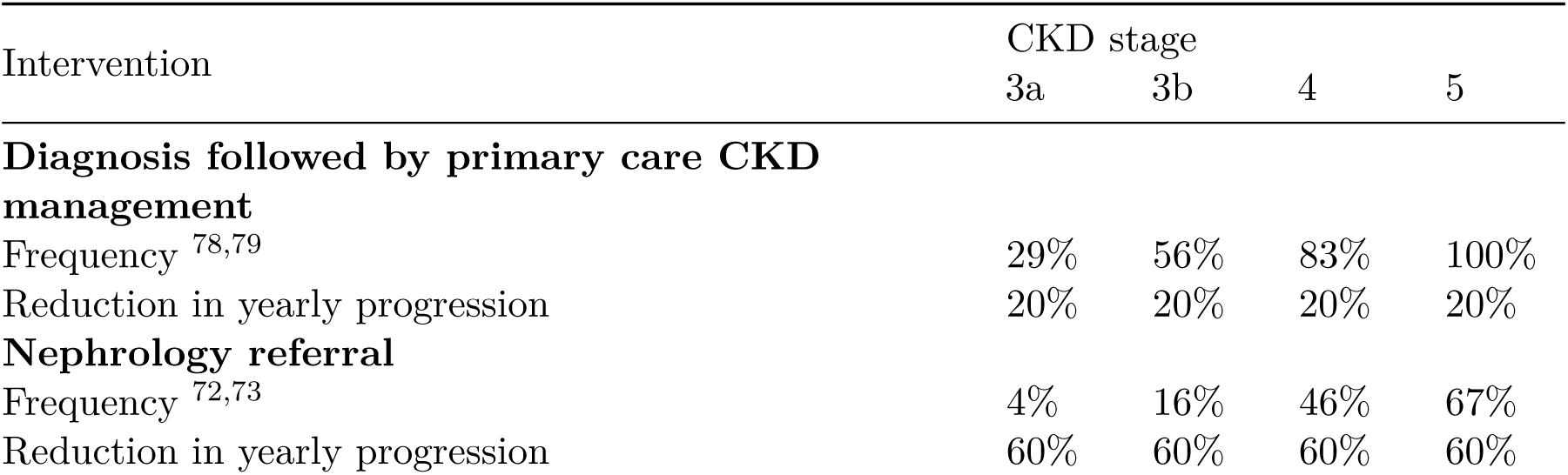
Frequency and effectiveness of interventions by chronic kidney disease (CKD) stage.^19,72,73,78–81^

| Intervention | CKD stage |  |  |  |
| --- | --- | --- | --- | --- |
|  | 3a | 3b | 4 | 5 |
| **Diagnosis followed by primary care CKD management** |  |  |  |  |
| Frequency <sup>78,79</sup> | 29% | 56% | 83% | 100% |
| Reduction in yearly progression | 20% | 20% | 20% | 20% |
| **Nephrology referral** |  |  |  |  |
| Frequency <sup>72,73</sup> | 4% | 16% | 46% | 67% |
| Reduction in yearly progression | 60% | 60% | 60% | 60% |

Calibration targets are included in Table S5, Table S6, Table S7, Table S8, Table S9. They correspond to the distribution of individuals across CKD stages, stratified by age groups and additional covariates.

**Table S5:** Number of people in the AFC cohort across chronic kidney disease (CKD) stages within age groups, stratified by binary sex. Cell counts at or below 11 were masked.

| Age | Stage 1 | Stage 2 | Stage 3a | Stage 3b | Stage 4 | Stage 5 |
| --- | --- | --- | --- | --- | --- | --- |
| Female |  |  |  |  |  |  |
| 30-39 | 25,178 | 4,930 | 148 | 28 | 15 | 15 |
| 40-49 | 38,234 | 14,241 | 624 | 141 | 50 | 40 |
| 50-59 | 44,174 | 38,129 | 3,091 | 630 | 169 | 88 |
| 60-69 | 38,250 | 55,072 | 8,888 | 2,526 | 520 | 139 |
| 70-79 | 15,597 | 41,234 | 14,266 | 4,704 | 883 | 154 |
| 80-89 | 841 | 17,069 | 8,313 | 4,485 | 929 | 59 |
| 90-99 | 11 | 2,046 | 1,627 | 1,122 | 277 | 11 |
| Male |  |  |  |  |  |  |
| 30-39 | 17,847 | 4,338 | 92 | 36 | 22 | 19 |
| 40-49 | 29,611 | 14,351 | 551 | 126 | 58 | 46 |
| 50-59 | 40,460 | 33,280 | 2,165 | 445 | 135 | 124 |
| 60-69 | 34,230 | 45,662 | 6,010 | 1,475 | 332 | 176 |
| 70-79 | 11,737 | 35,228 | 9,411 | 2,919 | 553 | 126 |
| 80-89 | 607 | 11,526 | 5,270 | 2,450 | 461 | 55 |
| 90-99 | 11 | 845 | 616 | 377 | 84 | 11 |

## S3: Sensitivity Analysis Supplement

We conducted a two-way sensitivity analysis to evaluate the impact of intervention parameters (frequency and effectiveness) on differences in expected survival. We considered the parameters described in Table S10.

All combinations of parameters that met the following criteria were considered. First, for each intervention type, we required the frequency of intervention at each consecutive stage to be higher or equal than that at the previous stage (unless both were at 100%). Second, at each stage, we required that the frequency of the nephrology intervention be less than or equal that of diagnosis. Finally, the effectiveness of nephrology was required to be higher than that of diagnosis alone. We examined all 72,226 parameter combinations that met these criteria and present the results below.

Increases in the frequency of interventions and in intervention effectiveness led to increases in additional life expectancy (comparing counterfactual and reference scenarios) for Black men and women, and decreases for non-Black men and women, in the general population (Figure S1, Figure S2) and among those who developed CKD in the reference scenario (Figure S3, Figure S4). Additional life expectancy was calculated across the whole life span from age 30. The magnitude of changes related to the diagnosis intervention did not surpass 1.7 months in the general population, and went up to 4.2 months among individuals who would reach CKD stage 3a in the reference scenario (if 100% of individuals reaching stage 3a would be immediately diagnosed and provided primary care interventions which would reduce their rate of progression by 40%). While magnitudes of the difference of life expectancy between scenarios were relatively small, the magnitude ^3^o^4^f reduction in life expectancy among non-Black individuals generally surpassed the magnitude of the improvement of life expectancy among Black individuals.

**Table S6:**
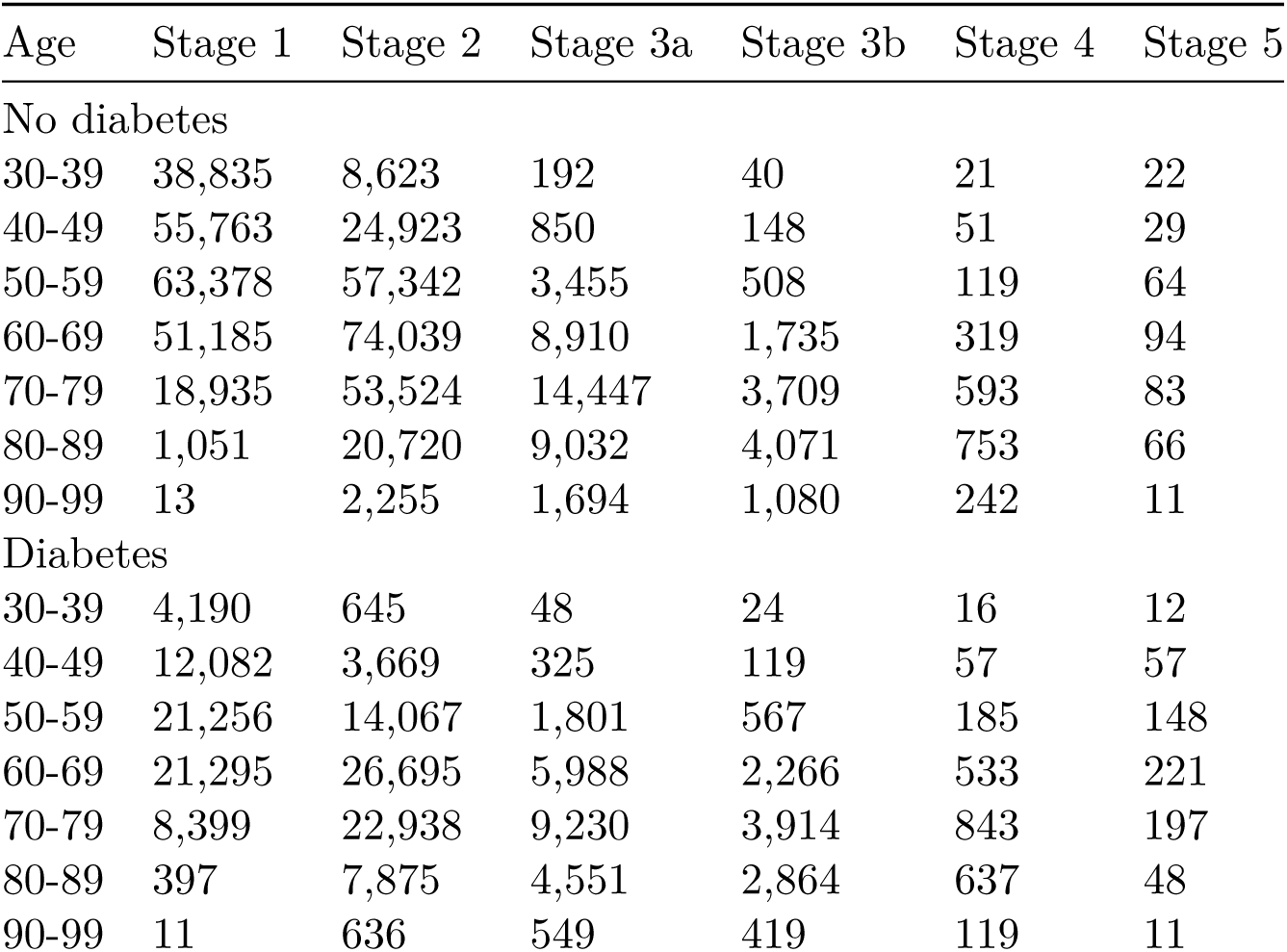
Number of people in the AFC cohort across chronic kidney disease (CKD) stages within age groups, stratified by diabetes status. Cell counts at or below 11 were masked.

## S4: Figures and Tables Supplement

**Table S7:** Number of people in the AFC cohort across chronic kidney disease (CKD) stages within age groups, stratified by hypertension status. Cell counts at or below 11 were masked.

| Age | Stage 1 | Stage 2 | Stage 3a | Stage 3b | Stage 4 | Stage 5 |
| --- | --- | --- | --- | --- | --- | --- |
| No hypertension |  |  |  |  |  |  |
| 30-39 | 32,132 | 6,706 | 124 | 16 | 11 | 11 |
| 40-49 | 39,967 | 16,441 | 423 | 48 | 13 | 11 |
| 50-59 | 37,751 | 31,784 | 1,485 | 152 | 33 | 27 |
| 60-69 | 24,968 | 32,604 | 2,714 | 441 | 88 | 40 |
| 70-79 | 7,107 | 17,728 | 3,298 | 681 | 102 | 22 |
| 80-89 | 311 | 5,380 | 1,733 | 547 | 100 | 11 |
| 90-99 | 11 | 508 | 280 | 131 | 42 | 11 |
| Hypertension |  |  |  |  |  |  |
| 30-39 | 10,893 | 2,562 | 116 | 48 | 31 | 25 |
| 40-49 | 27,878 | 12,151 | 752 | 219 | 95 | 78 |
| 50-59 | 46,883 | 39,625 | 3,771 | 923 | 271 | 185 |
| 60-69 | 47,512 | 68,130 | 12,184 | 3,560 | 764 | 275 |
| 70-79 | 20,227 | 58,734 | 20,379 | 6,942 | 1,334 | 258 |
| 80-89 | 1,137 | 23,215 | 11,850 | 6,388 | 1,290 | 103 |
| 90-99 | 16 | 2,383 | 1,963 | 1,368 | 319 | 14 |

**Table S8:** Number of people in the AFC cohort across chronic kidney disease (CKD) stages within age groups, stratified by ICE tertiles (from most to least deprived areas). Cell counts at or below 11 were masked.

| Age | Stage 1 | Stage 2 | Stage 3a | Stage 3b | Stage 4 | Stage 5 |
| --- | --- | --- | --- | --- | --- | --- |
| First tertile |  |  |  |  |  |  |
| 30-39 | 12,927 | 2,522 | 83 | 21 | 15 | 21 |
| 40-49 | 20,110 | 7,329 | 407 | 99 | 40 | 47 |
| 50-59 | 24,350 | 18,019 | 1,684 | 405 | 125 | 117 |
| 60-69 | 20,261 | 25,896 | 4,503 | 1,389 | 314 | 156 |
| 70-79 | 7,788 | 20,230 | 6,726 | 2,407 | 497 | 135 |
| 80-89 | 460 | 7,964 | 3,945 | 2,134 | 453 | 40 |
| 90-99 | 11 | 793 | 634 | 423 | 109 | 11 |
| Second tertile |  |  |  |  |  |  |
| 30-39 | 11,923 | 2,624 | 68 | 18 | 11 | 11 |
| 40-49 | 18,795 | 8,019 | 344 | 83 | 37 | 16 |
| 50-59 | 23,474 | 20,344 | 1,464 | 294 | 79 | 49 |
| 60-69 | 20,994 | 29,706 | 4,528 | 1,232 | 257 | 58 |
| 70-79 | 7,830 | 22,794 | 7,260 | 2,369 | 435 | 61 |
| 80-89 | 397 | 8,663 | 4,172 | 2,168 | 434 | 34 |
| 90-99 | 11 | 850 | 653 | 437 | 109 | 11 |
| Third tertile |  |  |  |  |  |  |
| 30-39 | 12,253 | 2,782 | 61 | 16 | 11 | 11 |
| 40-49 | 19,692 | 9,032 | 250 | 35 | 11 | 11 |
| 50-59 | 24,272 | 21,992 | 1,210 | 183 | 48 | 16 |
| 60-69 | 20,046 | 29,428 | 3,425 | 712 | 129 | 53 |
| 70-79 | 7,338 | 21,455 | 5,829 | 1,596 | 269 | 43 |
| 80-89 | 345 | 7,349 | 3,291 | 1,575 | 267 | 27 |
| 90-99 | 11 | 747 | 591 | 367 | 76 | 11 |

**Table S9:** Number of people in the AFC cohort across chronic kidney disease (CKD) stages within age groups, stratified by ICE tertiles (from most to least deprived areas). Cell counts at or below 11 were masked.

| Age | Stage 1 | Stage 2 | Stage 3a | Stage 3b | Stage 4 | Stage 5 |
| --- | --- | --- | --- | --- | --- | --- |
| First tertile |  |  |  |  |  |  |
| 30-39 | 13,026 | 2,565 | 81 | 23 | 14 | 16 |
| 40-49 | 20,069 | 7,492 | 392 | 101 | 45 | 44 |
| 50-59 | 23,925 | 18,337 | 1,720 | 393 | 123 | 111 |
| 60-69 | 20,458 | 26,717 | 4,697 | 1,436 | 342 | 153 |
| 70-79 | 7,892 | 20,917 | 7,029 | 2,468 | 520 | 134 |
| 80-89 | 486 | 8,555 | 4,228 | 2,285 | 482 | 46 |
| 90-99 | 11 | 870 | 702 | 494 | 129 | 11 |
| Second tertile |  |  |  |  |  |  |
| 30-39 | 12,068 | 2,685 | 81 | 15 | 13 | 11 |
| 40-49 | 18,699 | 8,125 | 345 | 74 | 30 | 20 |
| 50-59 | 23,673 | 20,410 | 1,429 | 300 | 74 | 45 |
| 60-69 | 20,357 | 29,070 | 4,177 | 1,140 | 226 | 61 |
| 70-79 | 7,677 | 22,220 | 6,916 | 2,258 | 403 | 61 |
| 80-89 | 381 | 8,321 | 4,064 | 2,064 | 399 | 27 |
| 90-99 | 11 | 835 | 635 | 408 | 95 | 11 |
| Third tertile |  |  |  |  |  |  |
| 30-39 | 12,009 | 2,678 | 50 | 17 | 11 | 11 |
| 40-49 | 19,829 | 8,763 | 264 | 42 | 12 | 11 |
| 50-59 | 24,498 | 21,608 | 1,209 | 189 | 55 | 26 |
| 60-69 | 20,486 | 29,243 | 3,582 | 757 | 132 | 53 |
| 70-79 | 7,387 | 21,342 | 5,870 | 1,646 | 278 | 44 |
| 80-89 | 335 | 7,100 | 3,116 | 1,528 | 273 | 28 |
| 90-99 | 11 | 685 | 541 | 325 | 70 | 11 |

**Table S10:** Parameters considered during sensitivity analysis. Effectiveness corresponds to multiplicative reduction of yearly eGFR decline speed.

| Stage | Original value | Sensitivity analysis values |
| --- | --- | --- |
| Diagnosis intervention frequency by stage |  |  |
| 3a | 0.29 | 0, 0.20, 0.40, 0.60, 0.80, 1.00 |
| 3b | 0.56 | 0.20, 0.40, 0.60, 0.80, 1.00 |
| 4 | 0.83 | 0.40, 0.60, 0.80, 1.00 |
| 5 | 1.00 | 0.90, 0.95, 1.00 |
| Nephrology intervention frequency by stage |  |  |
| 3a | 0.04 | 0, 0.05, 0.10 |
| 3b | 0.16 | 0, 0.10, 0.20 |
| 4 | 0.46 | 0.10, 0.30, 0.50, 0.70 |
| 5 | 0.67 | 0.40, 0.60, 0.80, 1.00 |
| Diagnosis effectiveness |  |  |
| all | 0.20 | 0.10, 0.20, 0.30, 0.40 |
| Nephrology effectiveness |  |  |
| all | 0.60 | 0.10, 0.30, 0.50, 0.70, 0.90 |

**Figure S1:**
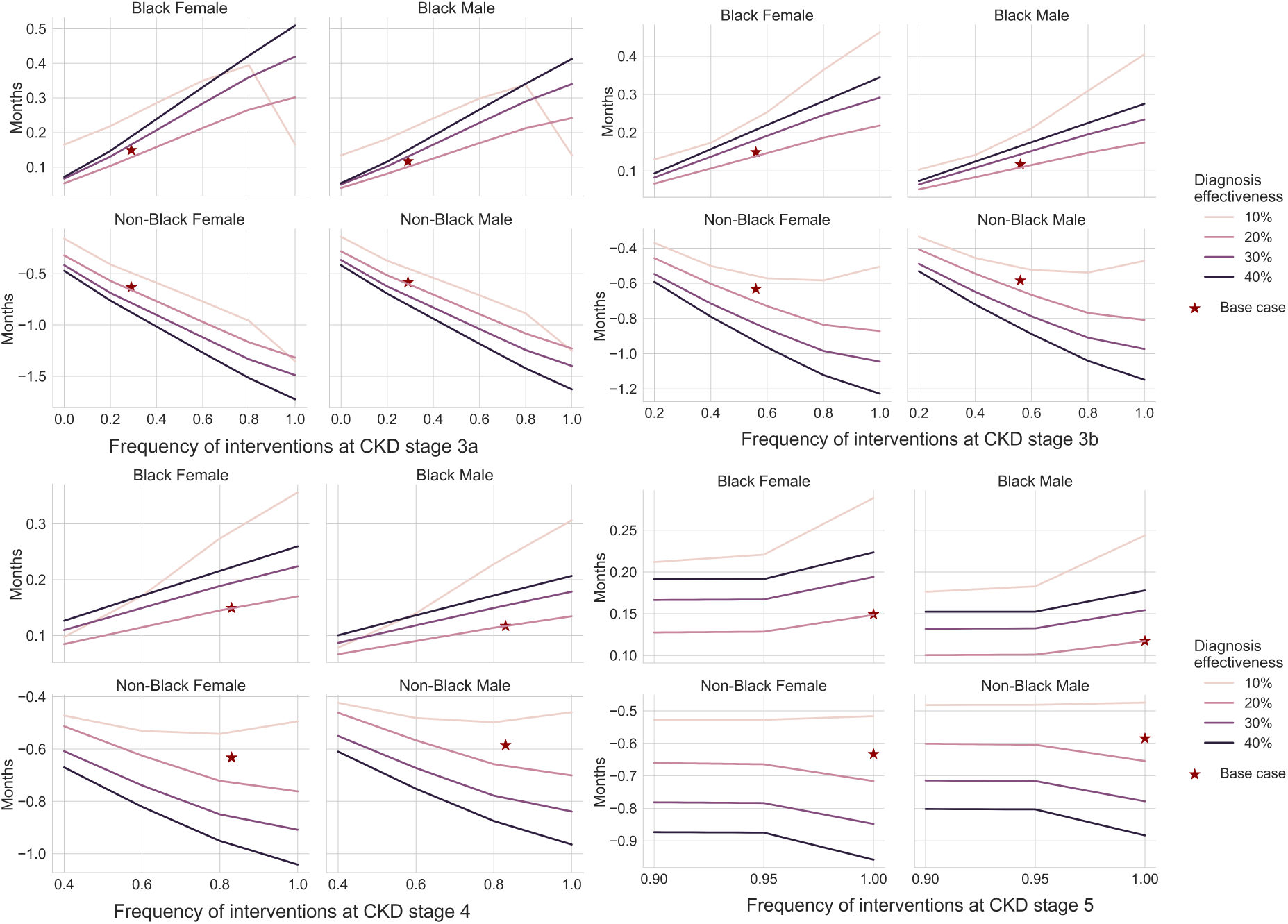
Mean additional months of life expectancy across different diagnosis frequency and effectiveness values.

**Figure S2:**
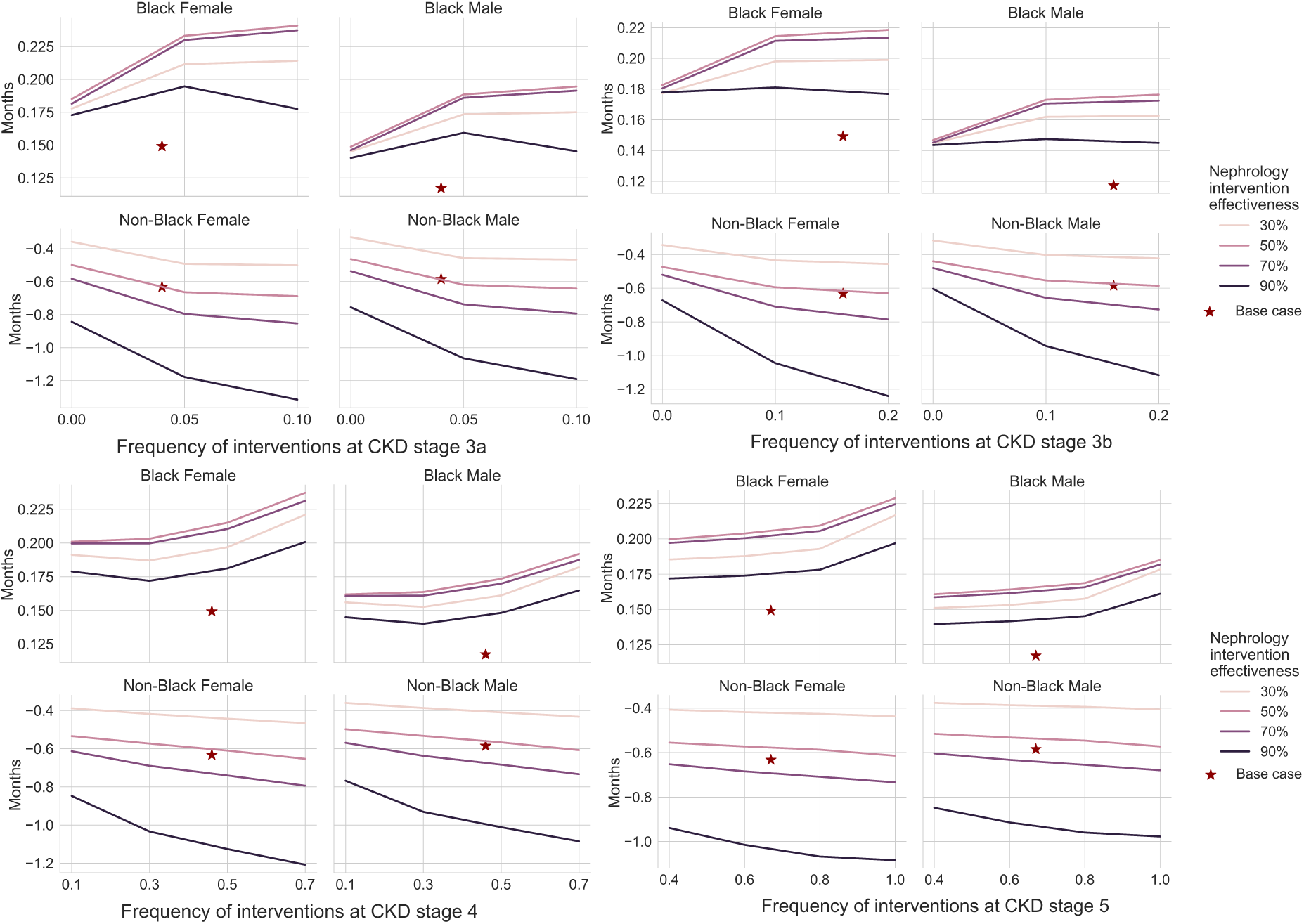
Mean additional months of life expectancy across different nephrology frequency and effectiveness values.

**Figure S3:**
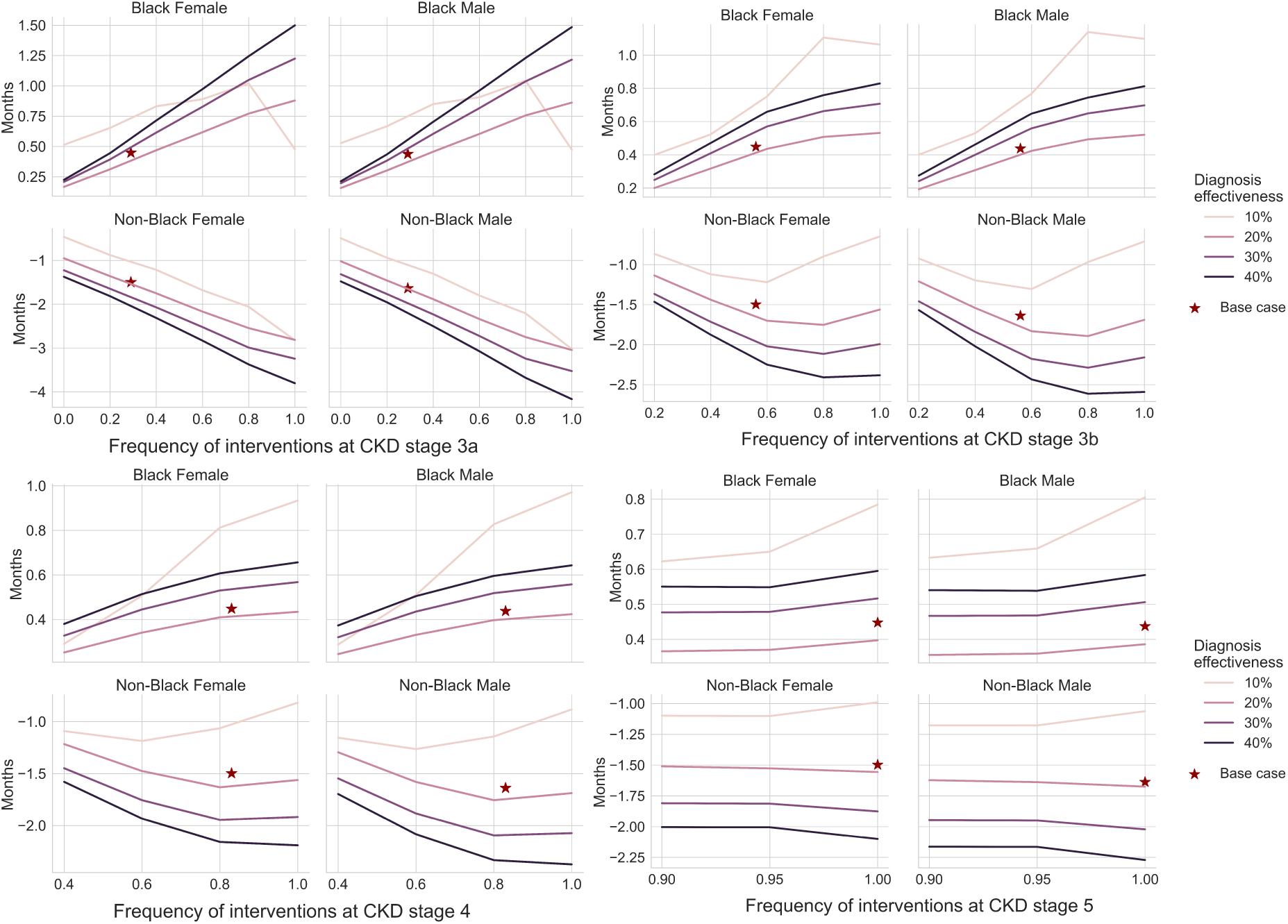
Mean additional months of life expectancy across different diagnosis frequency and effectiveness values among simulated individuals who reached CKD stage 3a in the reference scenario.

**Figure S4:**
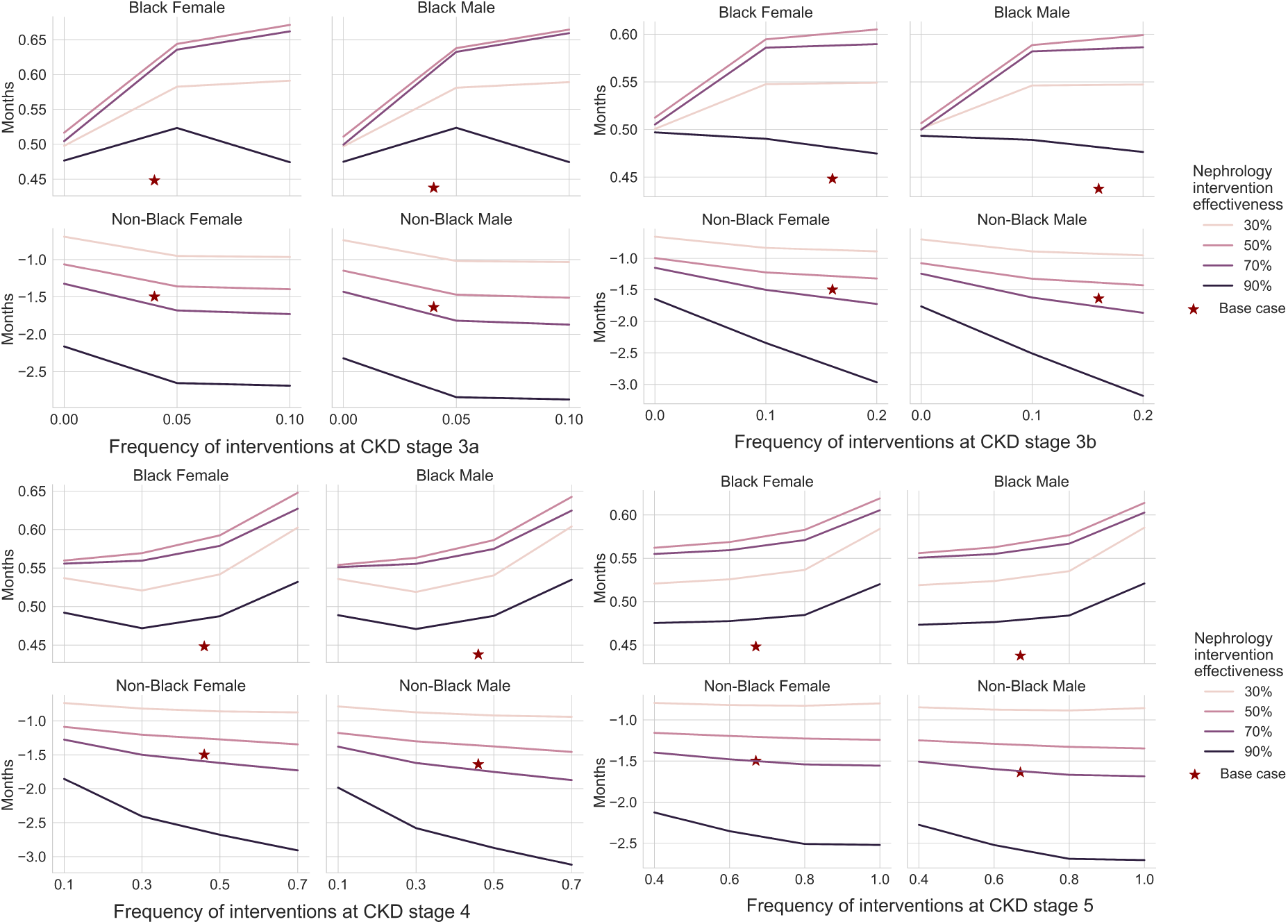
Mean additional months of life expectancy across different nephrology frequency and effectiveness values among simulated individuals who reached CKD stage 3a in the reference scenario.

**Figure S5:**
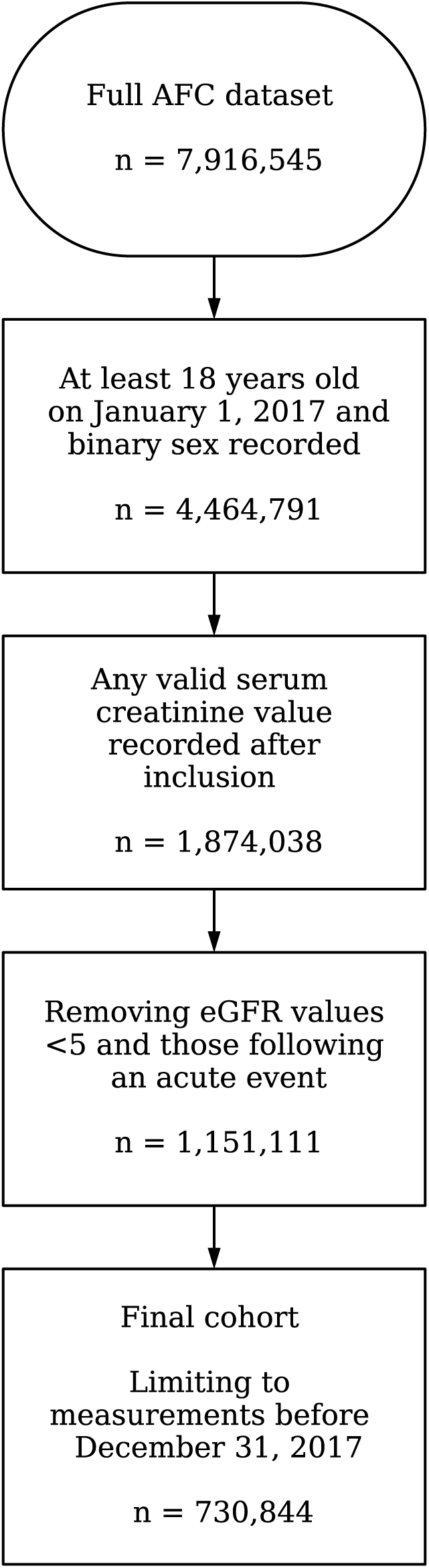
Cohort extraction flowchart.

**Figure S6:**
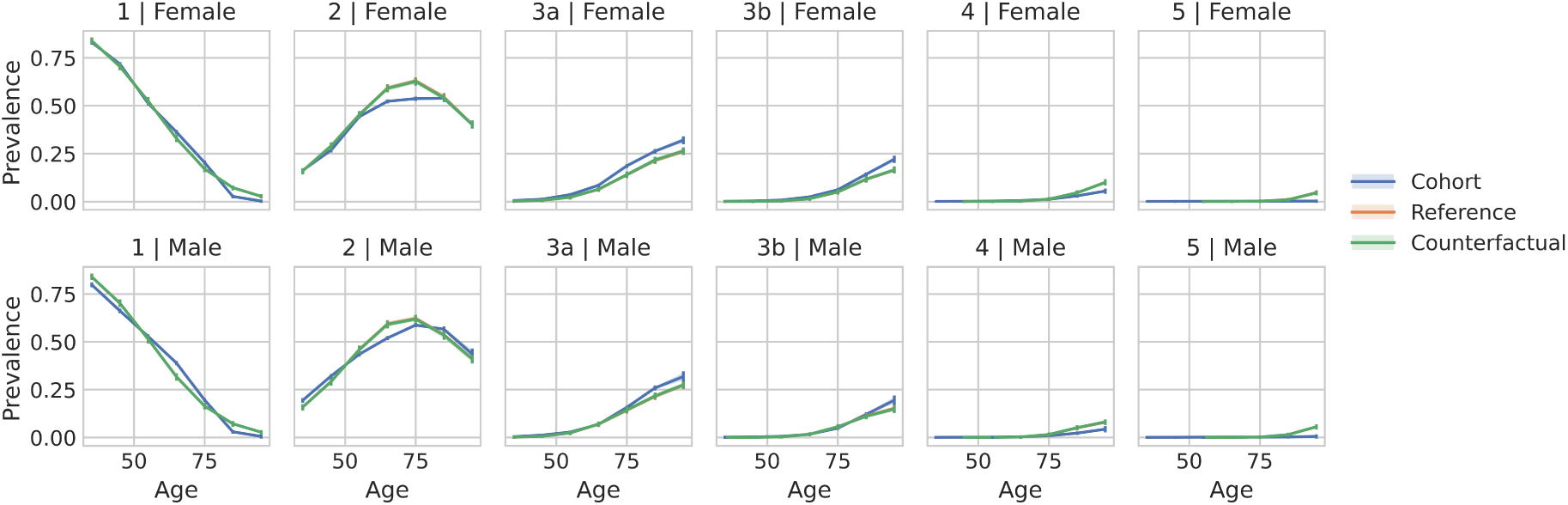
Stage prevalence across ages stratified by sex under two simulated scenarios (reference and counterfactual), compared to that observed in the AFC cohort.

**Figure S7:**
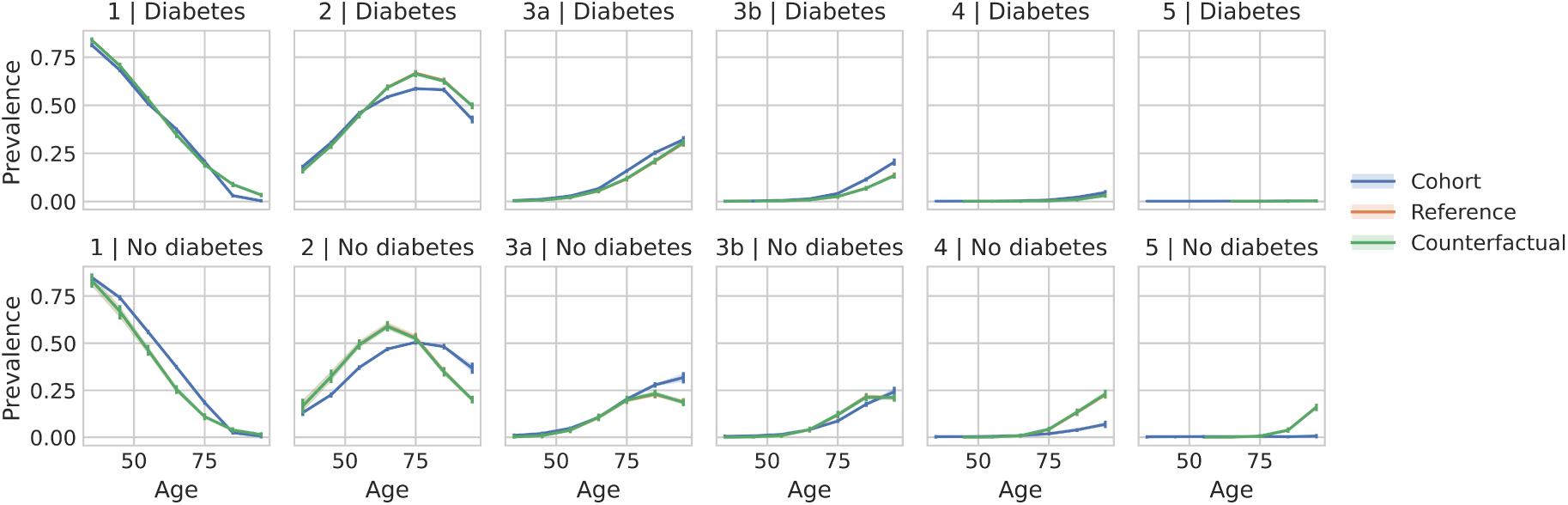
Stage prevalence across ages stratified by diabetes status under two simulated scenarios (reference and counterfactual), compared to that observed in the AFC cohort.

**Figure S8:**
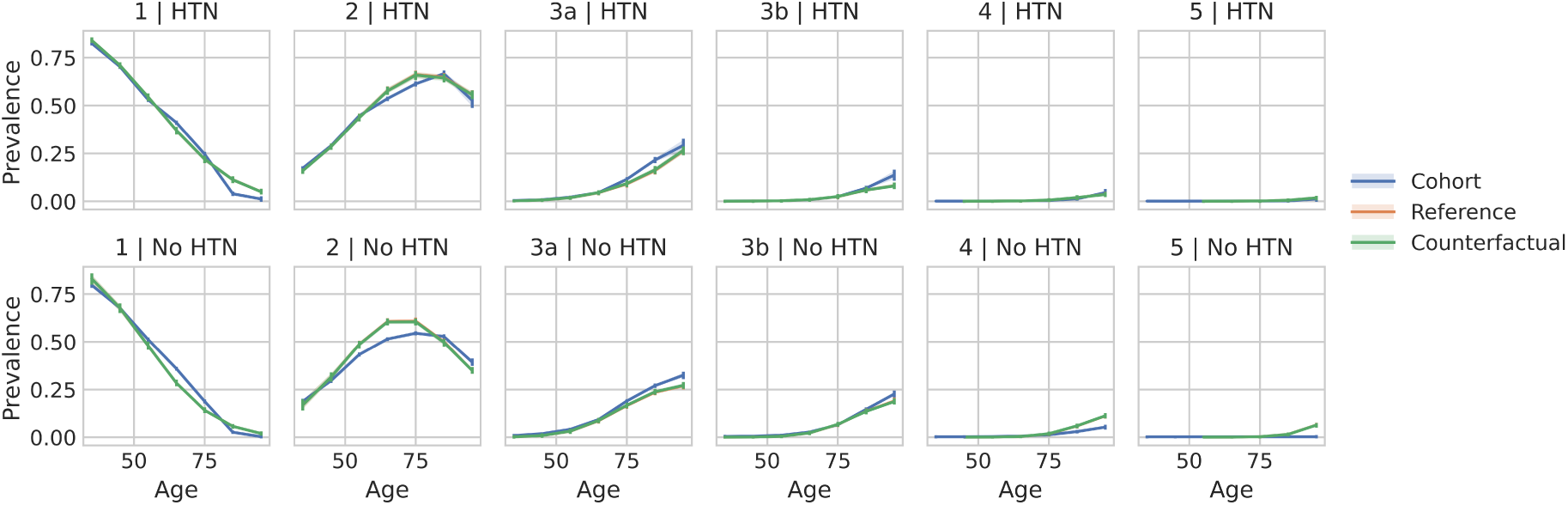
Stage prevalence across age stratified by hypertension (HTN) status under two simulated scenarios (reference and counterfactual), compared to that observed in the AFC cohort.

**Figure S9:**
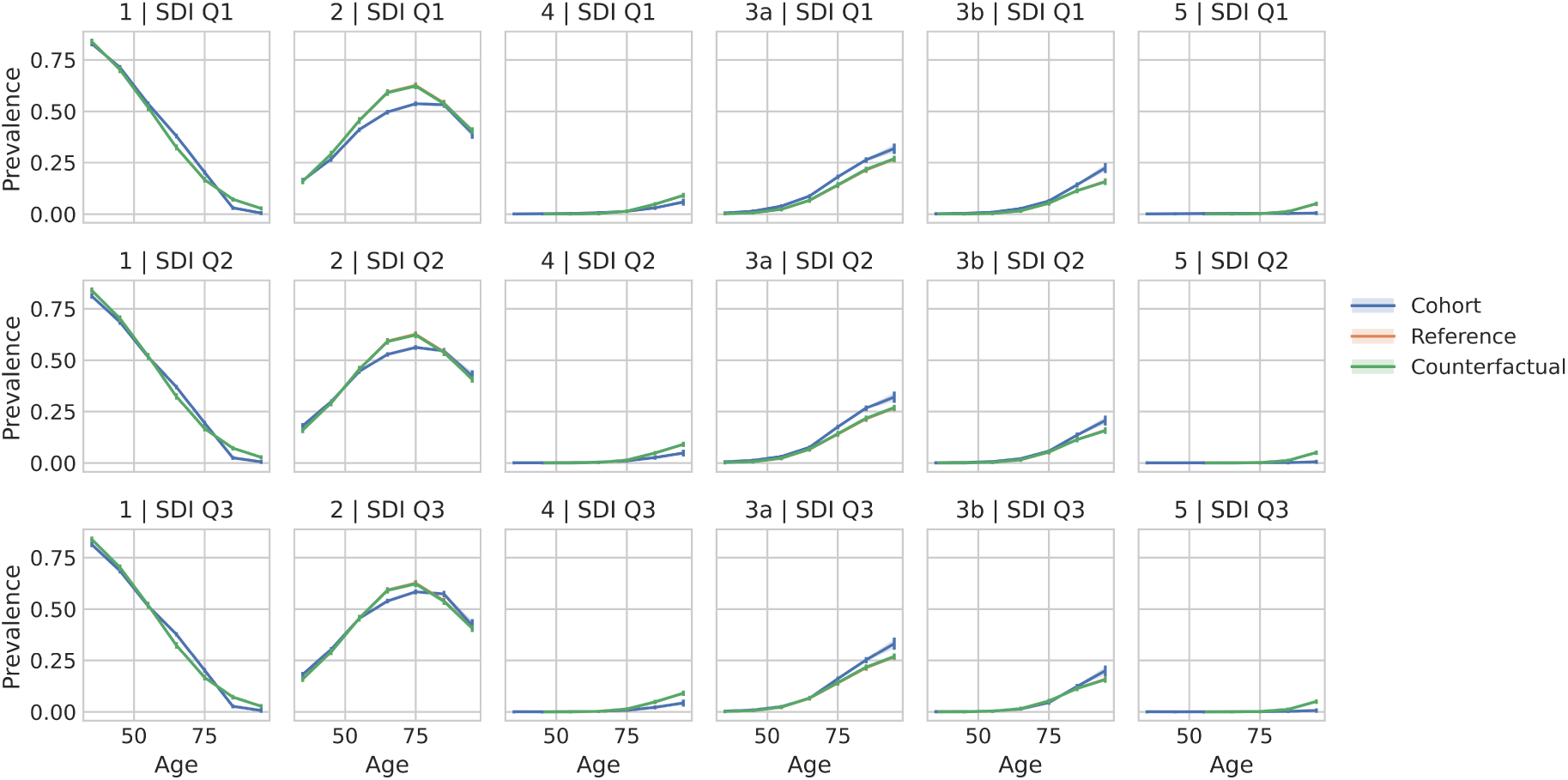
Stage prevalence across age stratified by SDI tertiles under two simulated scenarios (reference and counterfactual), compared to that observed in the AFC cohort. SDI tertiles range from most (Q1) to least deprived (Q3).

**Figure S10:**
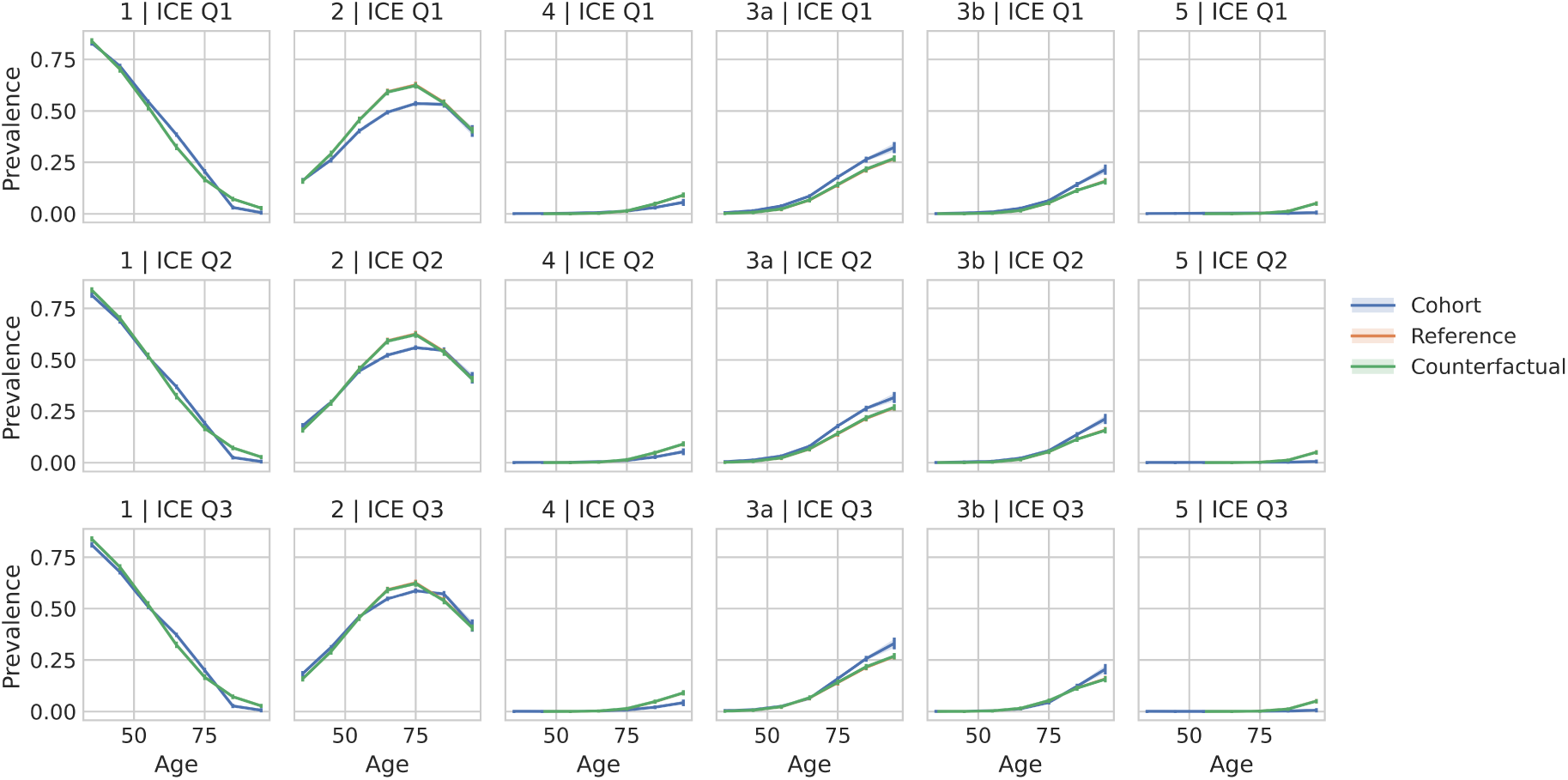
Stage prevalence across age stratified by ICE tertiles under two simulated scenarios (reference and counterfactual), compared to that observed in the AFC cohort. ICE tertiles range from most (Q1) to least deprived (Q3).

